# Safety, tolerability and clinical effects of BC007 (Rovunaptabin) on fatigue and quality of life in patients with post-COVID syndrome (reCOVer): a prospective, exploratory, randomised, placebo-controlled, double-blind, crossover phase IIa clinical trial

**DOI:** 10.1101/2024.12.13.24318856

**Authors:** Bettina Hohberger, Marion Ganslmayer, Thomas Harrer, Friedrich Kruse, Stefanie Maas, Tobias Borst, Ralph Heimke-Brinck, Andreas Stog, Thomas Knauer, Eva Rühl, Victoria Zeisberg, Adam Skornia, Alexander Bartsch, Armin Ströbel, Monika Wytopil, Carolin Merkel, Sophia Hofmann, Katja G. Schmidt, Petra Lakatos, Julia Schottenhamml, Martin Herrmann, Christian Mardin, Jürgen Rech

**Affiliations:** Department of Ophthalmology, Friedrich-Alexander-Universität Erlangen-Nürnberg and Uniklinikum Erlangen, Erlangen, Germany; Deutsches Zentrum für Immuntherapie (DZI), Friedrich-Alexander-Universität Erlangen-Nürnberg and Uniklinikum Erlangen; Department of Medicine 1, Friedrich-Alexander-Universität Erlangen-Nürnberg and Uniklinikum Erlangen, Erlangen, Germany; Infectious Diseases and Immunodeficiency Section, Department of Medicine 3 - Rheumatology and Immunology, Friedrich-Alexander-Universität Erlangen-Nürnberg and Uniklinikum Erlangen, Erlangen, Germany; Department of Medicine 3 - Rheumatology and Immunology, Friedrich-Alexander-Universität Erlangen-Nürnberg and Uniklinikum Erlangen, Erlangen, Germany; Center for Clinical Studies (CCS), Friedrich-Alexander-Universität Erlangen-Nürnberg and Uniklinikum Erlangen, Erlangen, Germany; Pharmacy of the University Hospital Erlangen, Uniklinikum Erlangen, Erlangen, Germany; Institute of Clinical and Molecular Virology, Friedrich-Alexander-Universität Erlangen-Nürnberg and Uniklinikum Erlangen, Erlangen, Germany; Department of Medicine 2, Friedrich-Alexander-Universität Erlangen-Nürnberg and Uniklinikum Erlangen, Erlangen, Germany

**Author notes:** contributed equally to first authorship (Bettina Hohberger, Marion Ganslmayer). contributed equally to senior authorship (Christian Mardin, Jürgen Rech).

**Keywords:** Post-COVID Syndrome, Long-COVID, BC007, PCS subtypes, GPCR, functional autoantibody, fatigue, quality of life

## Abstract

**Background:** As recent data suggest an involvement of GPCR-fAAb in PCS pathogenesis, neutralisation of such GPCR-fAAbs by BC007 could improve PCS symptoms. The aim of the reCOVer trial was to investigate safety, tolerability and clinical effects of BC007 on fatigue, its severity and quality of life in PCS patients.

**Methods:** reCOVer is a prospective, exploratory, randomised, placebo-controlled, double-blind, crossover phase IIa clinical trial with 1350 mg BC007 at the University of Erlangen-Nürnberg, Germany. Eligible participants were 18-80 years with GPCR-fAAb, whose PCS symptoms persisted ≥3 months after PCR-confirmed COVID-19, with fatigue as the major symptom (Bell score ≤60) and at least three of eight defined PCS symptoms. Participants were randomly assigned (1:1) according to a crossover design to either receive BC007 (sequence A) or placebo (sequence B) at day 0 and day 48 with a follow-up of 28 days, respectively. A crossover design was chosen to increase patient adherence. Occurrence of treatment-emergent adverse events (TEAEs) in comparison between sequence A and B from d0 to d28 and d0 to d70 were the primary and co-primary endpoint, respectively.

**Findings:** Between 31.10.2023 and 12.06.2024, 30 PCS patients were randomised and analysed. The trial has been concluded. Summarising all AE rates, no statistically significant differences between sequence A und sequence B were observed within day 28 and day 70. One report of a serious adverse event, not related to treatment, was recorded. As a secondary endpoint, BC007 showed a significant improvement on self-reported fatigue and its severity, as well as quality of life.

**Interpretation:** As BC007 was well tolerated and showed a significant improvement of fatigue and quality of life, it might offer a therapeutic option for an autoimmune subgroup of PCS patients.

**Trial registration:** EudraCT, number 2022-001781-35.

**Funding:** German Federal Ministry of Education and Research, German Research Foundation.

## Introduction

Post-COVID syndrome (PCS), also known as post-acute sequelae of SARS-CoV-2 (PASC), summarises clinical symptoms, persisting longer than 3 months after acute COVID-19 ^1^. Due to the complex and multifactorial pathogenesis, patients’ symptoms vary widely: next to long lasting fatigue with post-exertional malaise (PEM), e. g. headache, loss of concentration, hair loss, dyspnoea, chest pain, cognitive impairments (brain fog), myalgia, cardiac or gastrointestinal impairments were reported, partially not correlating with the severity of acute COVID-19 ^2^. Recent meta-analysis showed that prevalence of PCS is even up to 30% two years after COVID-19. Next to each individual burden, PCS has a remarkable impact on economy.

The exact pathogenesis of PCS is still elusive, yet different pathogenetic molecular pathways, indicating diverse PCS subgroups, are proposed: *long-term organ dysfunction* by virus and inflammation induced organ damage (e.g. pneumonia, myocarditis) and incomplete or aberrant repair (e.g. lung fibrosis); *reactivation of viral pathogens* (e.g. herpesviruses) or *viral (SARS-CoV-2) persistence,* inducing chronic inflammation with ongoing immune activation ^3^; *endothelial dysfunction* and *clotting phenomena*: combination of microthrombosis (i.e. microclotting), NETosis, an altered blood rheology and/or endothelial dysfunction might result in long-term ischemia-reperfusion damage ^4–6^; *autonomic nervous system dysregulation*, triggering e.g. cardiovascular autonomic dysfunction (CVAD, including the postural orthostatic tachycardia syndrome, POTS); *immune dysregulation and autoimmunity* based on viral attack with generation of different autoantibodies, partially with functional activity ^6^. Understanding these mechanisms and their overlapping, hence resulting in the necessity of patient stratification, are crucial for developing effective and causal treatments for patients’ care.

One of the proposed supgroup of patients with PCS exhibit different pattern of functional autoantibodies targeting G-protein coupled receptors (GPCR-fAAb), showing a link to an impaired capillary microcirculation. Neutralisation of those GPCR-fAAb by BC007 (Rovunaptabin, Berlin Cures GmbH, Berlin, Germany) was accompanied by an improvement of capillary microcirculation and patient’s PCS symptoms in an individual healing attempt ^7^. BC007 is a 15-mer single strand DNA oligonucleotide, thrombin binding aptamer (TBA)^8^, later shown to bind and neutralise various GPCR-fAAb *in-vitro* and *in-vivo* ^9^. Clinical data of a phase I study (NCT02955420) showed a good safety profile in healthy participants with a mild-to-moderate increase in activated Partial Thromboplastin Time (aPTT) ^10^. Next to its primary indication as an anticoagulant, BC007 was tested in a first clinical phase IIa study in patients with dilated cardiomyopathy as GPCR-fAAb neutralisation agent (NCT04192214). We conducted a prospective, exploratory, randomised, placebo-controlled, double-blinded, crossover phase IIa clinical trial of BC007 in an autoimmune subgroup of patients with PCS (reCOVer). Here we report on safety, tolerability and clinical effects of BC007 on fatigue and quality of life in patients with PCS.

## Methods

### Trial design

The prospective, randomised, controlled, double-blind cross-over phase IIa clinical trial of BC-007 (reCOVer) was conducted at the Department of Ophthalmology, University of Erlangen-Nürnberg (UKER), Erlangen, Germany as an investigator initiated trial (IIT). A total of 37 patients with PCS were selected for screening according to the inclusion and exclusion criteria. A total of 30 patients were included in the study and randomised in a 1:1 ratio. The duration of participation was 91 days per participant according study protocol. The crossover design was chosen due to increase of patients’ adherence to overall study participation (i.e. each participant will receive BC007). As no causal therapy is available in PCS treatment, no specific washout phase was necessary prior to randomisation (comorbidities can be seen in suppl. Table 1 and 2, concurrent medication in suppl. table 3. For this initial exploratory pilot study, a sample size of n=30 was considered sufficient, to obtain data for a subsequent study with an effect size of 0.35 ^11^. The clinical trial was approved by German Competent Authority (Federal Institute for Drugs and Medical Devices, BfArM) and the ethics committee of the Friedrich-Alexander University Erlangen-Nürnberg (23-50-Az). It is registered in the European Union Drug Regulating Authorities Clinical Trials Database (EudraCT No.: 2022-001781-35). Prior to enrolment the trial was registered in the German Clinical Trials Register (DRKS00032237) which also provides the protocol. The clinical trial was conducted in accordance with the study protocol, regulatory requirements, the ICH-GCP and the ethical principles of the Declaration of Helsinki. Written informed consent was obtained from all participants in the study. The clinical trial has been completed within 12 months on September 2024.

### Participants

Adult patients, of all genders between ages of 18-80 years were eligible for inclusion. Previous SARS-CoV-2 infection had to be confirmed by a positive polymerase chain reaction (PCR) test. PCS symptoms had to persist at least 3 months prior to study inclusion, with a clinically relevant fatigue (Bell score ≤60) and at least three of the following symptoms: Exercise intolerance, exertional dyspnoea, concentration disorders, brain fog, fatigue, unsteady gait, disorders of the sense of taste and/or smell, headaches. Furthermore, the presence of GPCR-fAAb was a prerequisite for participation. The therapeutic use of anticoagulants, that could interact with the investigational medicinal product BC007 (IMP) and increase the tendency to bleed, was prohibited before and during the study for patient safety. All participants showed no signs of long-term COVID-19 induced organ damage. Patients with obstructive sleep apnoea syndrome (OSAS), obesity with a BMI >30 kg/m², acute and severe organ damage to the heart and lungs, malignancies or remission of malignant diseases within the last 2 years were excluded. No intake of interfering drugs that could lead to fatigue (e. g. opiates, antihistamines, antidepressants, NMDA antagonists) was permitted. Furthermore, eye diseases with impairment of the retinal microcirculation, systemic diseases with eye involvement, glaucoma, pregnancy and breastfeeding were excluded.

### Randomisation and masking

The randomisation sequence was generated by the statistician before the start of the enrolment with the maximal procedure with software R 4.3.0, a 1:1 ratio and a maximum tolerated imbalance of 6 ^12^. The length of the randomisation sequence was 40 to account for possible screening failures. The study was performed in a double-blinded manner, where participants and investigators were blinded. Unblinded members of the clinical trial team packed the drug and performed statistical analysis. The blinding was not broken during the trial, as no medical emergency occurred. Unblinded persons of the pharmacy, being responsible for allocation, produced the ready-to-use infusion of BC007 for intravenous application. Allocation concealment was done by pharmacy of UKER according to the randomisation list. Consequently, 1350 mg BC007 (in 50 ml NaCl 0·9%) or placebo were prepared and labelled in accordance with regulatory requirements. The carrier solution of 50 ml 0·9% sodium chloride, which had the same appearance as BC007, was used as placebo.

### Procedures

Eligible participants entered the randomised, double-blind, crossover treatment periods (sequence A and sequence B), receiving either 1350 mg BC007 or placebo at day 0 (V2) with a crossover at day 42 (V8). The allocation was generated by the Center for Clinical Studies CCS (AStr), participant enrolment was done by the investigator (AS, TK, BH, VZ, ASk). Assignment to sequence was done by the pharmacy. Sequence A received 1350 mg BC007 followed by placebo, sequence B received placebo, followed by 1350 mg BC007. Total study duration per participants was 91 days, including 14 visits: After screening (V1), first study treatment with either IMP or placebo was administered (V2, day 0) with follow-up visits at the next two days respectively (V3, V4). Further study visits were after 1 week (V5), 2 weeks (V6) and 4 weeks (V7). After a 14-day break the switch to the second treatment with IMP or placebo in the crossover procedure began on Day 42 (V8). The study visits were conducted on day 43 (V9), day 44 (V10), day 49 (V11), day 56 (V12), day 70 (V13) and day 91 (V14 [EOS]). Prior to each treatment, the presence of a negative SARS-COV-2 antigen test, not older than 24 hours, was required. Treatment was done by intravenous application of 1350 mg BC007 (in 50 ml NaCl 0·9%) within 75-90 min or placebo (50ml NaCl 0·9% alone) within 75-90 min.

### Safety

For analysis of safety and tolerability adverse events (AE) and vital signs were recorded during the whole observation period. Haematology, blood-chemistry and coagulation tests were performed at all study visits. A physical examination was done on both treatment days (day 0/42), day 1, day 28, day 43 and day 70. GPCR-fAAb testing was done at screening, day 7, day 28, day 49 and day 70, respectively. Fatigue scores and quality of life were assessed at all visits from day 0 to day 91 (except at V1, V9). 6 minute walk test (6MWT) was performed at day 0, day 7, day 28, day 42, day 49, day 70, and day 91. Occurrence of adverse events (AEs), adverse reaction (ARs), serious adverse event (SAEs) and serious adverse reaction (SARs) was recorded during the entire observation period, classified according to the Common Terminology Criteria for Adverse Events CTCAE (5 categories: mild-moderate-severe-life-threatening-fatal and coded using MedDRA v. 27.0. Serious adverse events were reported according to regulatory requirements. All relevant data was recorded in the eCRF.

### Outcomes

Primary endpoint was the occurrence of treatment-emergent adverse events (TEAEs) in the period from V2 (after administration of the investigational product) to V7 in the comparison between sequence A and sequence B. Coprimary endpoint was the occurrence of treatment-related adverse events (TEAEs) in the period from V2 (after administration of the investigational product) to V13 in the comparison between sequence A and sequence B.

The treatment effect of BC007 on self-reported fatigue and self-reported quality of life was assessed by the following second endpoints: the Functional Assessment of Chronic Illness Therapy (FACIT) Fatigue Scale and Chalder Fatigue Scale (for fatigue), the Bell scale and Fatigue Severity Scale (FSS, for severity of fatigue), the Canadian Criteria (CCC, for the presence of myalgic encephalomyelitis/chronic fatigue syndrome (ME/CFS)), SF-36 questionnaire (for quality of life), and 6MWT (Borg scales). The FACIT-Fatigue Scale is a self-reported 13-item questionnaire, assessing fatigue and its impact on daily activities over the past seven days. It is rated on a four-point Likert scale (4 = „not tired at all” to 0 = „very tired”). Rating is scored from 0 to 52 (healthy). The 11-item Chalder Fatigue Scale (CFS) assesses physical fatigue (questions 1-7) and mental fatigue (questions 8-11), scored with 4 response options ranging from ‘less’ to ‘much more’. Rating is scored from 0 (healthy) to 33. The Bell Scale assess fatigue severity, comprising a total of 10 responses with a rating from 0 to 100 (healthy). The 9-item FSS is scored from 1 („disagree“) to 7 („completely agree“). The lower scores, the healthier the participant is. CCC comprises 7 categories, including fatigue, PEM, sleep dysfunction, pain neurological and cognitive, autonomic, neuroendocrine and immune manifestations for a binary diagnosis of ME/CFS. The 5-item DePaul Symptom Questionnaire -Post-Exertional Malaise short form 8 (DSQ-PEM) is scored from 0 (not present) to severe (including grading from 1 (mild) to 4 (very severe); additional 5 items were recorded in a binary and time-coded score.

Quality of life was assessed by the 36-item Short Form (SF-36), including 8 subscales, the physical component summary (PCS) and the mental component summary (MCS). Rating is scored from 0 to 100 (healthy). 6MWT registers walking distance in a self-determined speed within 6 minutes, missing data for persons, which were unable to perform the 6MWT, were replaced by the value 0. BORG rating of perceived exertion scale assessed the subjectively perceived exertion during 6MWT, graded from 6 (no exertion) to 20 (maximum exertion). BORG-dyspnoe-scala assessed the subjectively perceived dyspnoea with a scoring from 0 (no) to 10 (maximum).

### Cardiomyocyte bioassay

For the detection of GPCR-fAAb, a cardiomyocyte (CM) bioassay was performed by G. Wallukat (Berlin Cures GmbH) as described previously^13^. IgGs from patient sera were heat inactived (56°C) and dialysed (dialysis buffer, a 0·15 M NaCl, 10 mM phosphate buffer, pH 7·4; Membra-Cel MD 44, 14 kDa, Serva, Heidelberg, Germany) and stored at -20°C. CMs were isolated from new-born Wistar rats and maintained in culture for maximum 8 days. IgG fraction was added to the cells in final dilution of 1:50 for 60 min following the determination of basal cell beating. Depending on the direction of the Δfrequency: positive chronotropic GPCR-fAAb (β2-fAAb, AT1-fAAb, α1-fAAb, Noci-fAAb) were identified with following inhibitors: ICI118,551 (0·1 μM), losartan (1 μM), urapidil (1 μM) and J113397-7 (0·1 μM), respectively; to determine negative chronotropic GPCR-fAAb (MAS-AAb, M2-AAb, ETA-AAb) following inhibitors were used: A779 (1 μM), atropine (1 µM) and BQ123 (0·1 μM). All inhibitors were from Sigma-Aldrich Chemie GmbH (Taufkirchen, Germany).

### Statistical Analysis

The statistical analysis was pre-specified in a statistical analysis plan, which was finalised before the start of the enrolment. The statistical analysis was based on a frequentist approach; significance level was set to 5% and tests and confidence intervals were two-sided. The primary variable was the relative frequency of patients with at least one TEAE in the period from baseline to day 28. This frequency is calculated for both sequences A and B and a 95%-confidence interval for the difference of these two frequencies is estimated ^14^. Also, the rate of TEAE per day was calculated and compared between the two sequences by a Poisson-test. The same methods were applied to the Coprimary variable (TEAE from baseline to day 70).

For all other variables the data analysis was split in two parts: data acquired before the cross-over at visit 8 and for all data up to visit 13.

When considering only the data before the crossover, the study design is that of a parallel group trial with a baseline (visit 2) and five repeated measurements after intervention. Methods have been described previously were applied ^15^ ^16^. Equation for the linear model was:

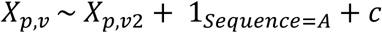

where p is the number of the patient in 1, …30, v is the visit in 3, …7, Xp,v is the value of a variable of patient p at visit v, 1Sequence=A is the indicator function for sequence A and c is a constant. In this model the coefficient for the indicator function is an estimator for the treatment effect. The treatment effect was tested for ≠ 0.

Essentially, the mean of all post-baseline-measurements was calculated, adjusted for the baseline-measurement and then compared between the two sequences.

If the baseline-value of a patient of a variable was missing this patient was omitted from this analysis of this variable. A missing value after baseline did no lead to an omission of the patient, when at least one post-baseline-measurement was available. Missing values were not imputed.

When considering all data, methods for cross-over studies described by Senn *et al* ^17^, section 3 and 4 were applied. A linear model similar to the one above was applied to estimate the cross-over difference (assuming no carry over or period effects):

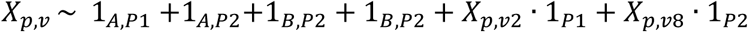

The contrast to estimate the cross over difference was:

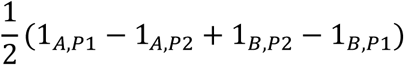

This contrast was tested for≠ 0. Additionally, both models were also estimated without the adjustment for the baseline-values.

### Role of funding source

The funders of the study (German Federal Ministry of Education and Research, BMBF, 01EP2108A, reCOVer; 01EO2105, iIMMUNE_ACS; German Research Foundation, DFG; 401821119/GRK2504) had no role in study design and conduct, data collection, analysis and interpretation, as well as writing of the manuscript.

## Results

Screening was carried out within 217 days (between 21.11.2023 and 25.06.2024); 37 patients with PCS were screened and assessed for eligibility, of whom 30 could be recruited for participation (figure 1). Seven persons were excluded because they declined participation and withdrew consent (n=4), did not fulfil either the inclusion or exclusion criteria due to facts after screening (n=1), noncompliance (n=1) or due to a seronegativity for GPCR-fAAb (n=1). Last patient out was on 10.09.2024.

**Figure 1:**
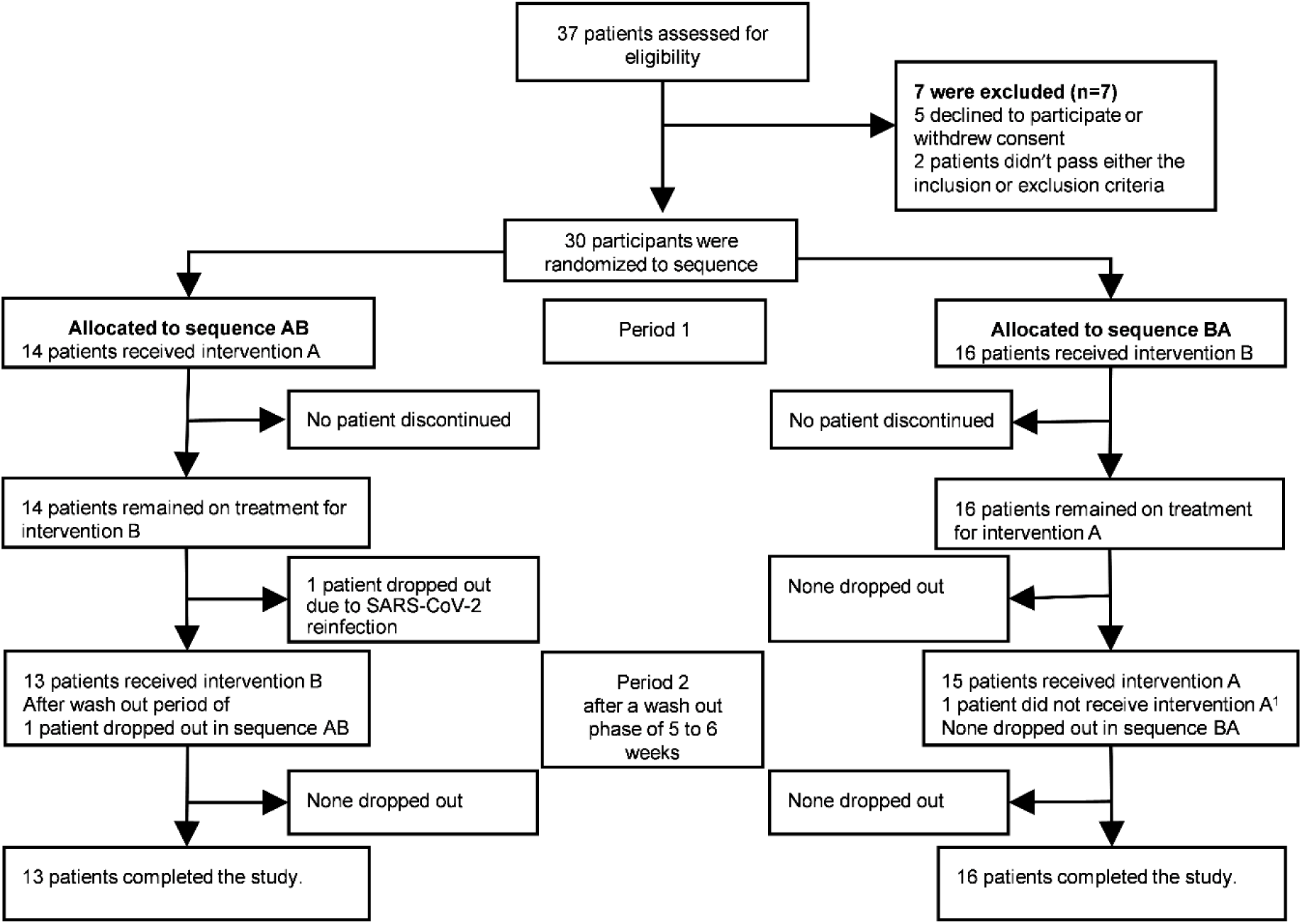
Trial profile. The safety and dosage population included all treated participants who took at least one administration of study drug. The population enrolled was randomised by our study pharmacy of the Universitätsklinikum Erlangen. Randomised participants passed all study inclusion or exclusion criteria. Before the infusion of either placebo or BC007 (dosage: 1350 mg) patients had to undergo a gargle water PCR-COVID-test. Patients received either Verum/Placebo, i.e. sequence A/B, on their 2nd or 8th visit. In total 14 visits were planned for each study-participant. The crossover design was divided in two periods seven with visits each for both study Arms A/B respectively. Missing visits in sequence A n=3. Missing visits in sequence B n=2.Total missing visits in both sequences A and B n=5. Sequence AB = verum followed by placebo, Sequence BA = placebo followed by verum; ^1^due to medical reasons (allergic reaction after first infusion).

Thirty participants were randomly assigned to the prospective double-blind cross-over treatment (Figure 2): sequence A (BC007 ➔ placebo; n=14), sequence B (placebo ➔ BC007; n=16). One participant, assigned to sequence A, was withdrawn due to acute COVID-19 infection (i. e. exclusion criteria, drop out) after day 28. Twenty-nine participants completed the trial. Of all participants’ visits, five visits of three participants were not realised due to the patients’ malaise (n=3) or illness (n=2). Therefore, thirty participants completed V7 including complete data set for primary outcome.

**Figure 2:**
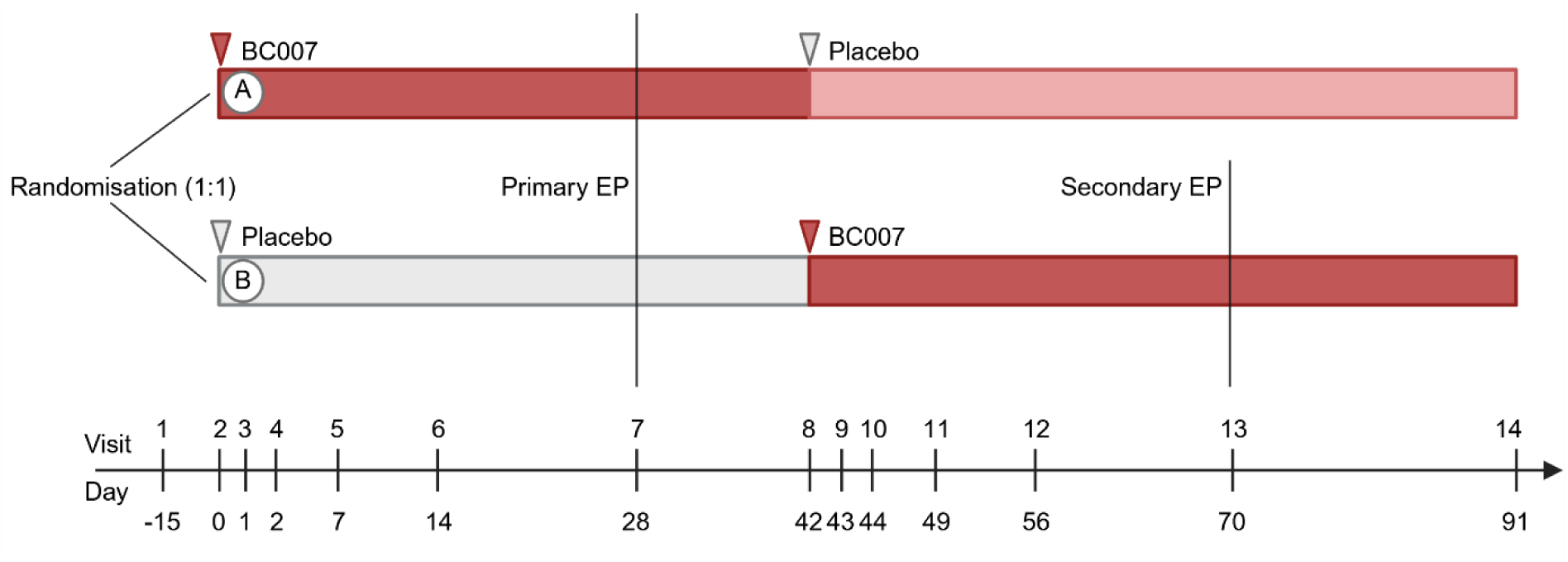
Overview of visit schedule and cross-over design. EP: Endpoint.

Demographic data can be seen in table 1, comorbidities are listed in supplementary table 1 and 2 and co-medications in supplementary table 3. Information regarding COVID-19 vaccination can be found in supplementary table 4. No clinically meaningful differences were observed between participants of the sequence groups.

**Table 1:** Demographic data.

|  | Sequence A (14) | Sequence B (16) | Total 30 |
| --- | --- | --- | --- |
| Age [years] | 41.0 ± 10.44 | 39.0 ± 7.92 | 39.9 ± 9.08 |
| Duration of PCS [days] | 1136 ± 283 | 1061 ± 232 | 1096 ± 256 |
| Male | 10 (71.4%) | 10 (62.5%) | 20 (66.7%) |
| Female | 4 (28.6%) | 6 (37.5%) | 10 (33.3%) |
| Smoker | 0 (0.0%) | 0 (0.0%) | 0 (0.0%) |
| Not smoking | 8 (57.1%) | 10 (62.5%) | 18 (60.0%) |
| Previous smoker | 6 (42.9%) | 6 (37.5%) | 12 (40.0%) |

PCS: Post-COVID Syndrome, NaN: Not a number. Age and Duration of PCS are given as means with SD.

The study primary safety endpoint was the occurrence of therapy-related adverse events (TEAEs) in the period from V2 (after 1st treatment) to V7 (day 28) between sequence A and sequence B. The number of participants with at least one TEAE from baseline to day 28 was six of 14 in sequence A (42·86%) and three of 16 in sequence B (18·75%), respectively. The confidence interval (CI) of the difference between both sequences was -14·80%; 63·02%. Comparing these AE rates between sequence A and sequence B, no statistical difference was observed (p=0·1299, Poisson-test). The ratio of the two event rates (AE/time) was 2·286 with a 95% CI of [0·71; 8·52].

Analysis of the co-primary endpoint (i.e. TAE from V2 until day 70) yielded that eight of 14 in sequence A (57·14%) and nine of 16 in sequence B (56·25%) participants reported at least one TEAE from baseline to day 70, which was not significantly different (p=0·1352, CI -35·54%; 37·32%, Poisson test). The ratio of the two event rates was 1·87 with a 95% CI of [0·84; 4·38]. Summarising all AE rates, no statistically significant differences between sequence A und sequence B were observed (p=0·1060; Poisson test). The ratio of the two event rates was 1·839 with a 95% CI of 0·85; 4·16. During treatment no clinically significant changes in vital signs were observed. One report of a serious adverse event, not related to treatment, was recorded. Overall, BC007 was well tolerated.

In line with its described anticoagulatory effects, BC007 significantly increased aPTT 2h after infusion in Sequence B, but not Sequence A (Wilcoxon signed-rank test, p=0.0026, Suppl. Table 5, Suppl. Figure 1). A significant increase in INR was observed in sequence A 2h after placebo infusion (Wilcoxon signed-rank test, p=0.0010, Suppl. Table 6, Suppl. Figure 1). No effect on the Quick value was seen in either sequence (Suppl. Table 7), however it was not evaluated 2h after infusion. A summary of all reported adverse events can be seen in supplementary table 8 and 9. Treatment effects of BC007 on fatigue, severity of fatigue and subjective rating of ME/CFS is presented in table 2.

**Table 2:** Treatment effect of BC007.

| Until day 28 | Treatment effect with baseline | 95% CI | SD | p-value | t-value |
| --- | --- | --- | --- | --- | --- |
| FACIT | 2.91 | 0.17 - 5.65 | 1.38 | 0.0378 | 2.10 |
| Bell | 7.54 | 3.43 - 11.64 | 2.07 | 0.0004 | 3.64 |
| FSS | -0.43 | -0.75 - -0.11 | 0.16 | 0.0088 | -2.66 |
| SF36 |  |  |  |  |  |
| Vitality | 6.65 | 3.38 - 9.93 | 1.65 | 0.0001 | 2.97 |
| General health | 4.86 | 1.04 - 8.67 | 1.92 | 0.0130 | 2.52 |
| Social Functioning | 6.46 | 0.31 - 12.61 | 3.10 | 0.0396 | 2.08 |
| Mental component | 2.73 | 0.19 - 5.28 | 1.28 | 0.0357 | 2.13 |
| **Crossover** |  |  |  |  |  |
| FACIT | 2.73 | 0.92 - 4.54 | 0.92 | 0.0033 | 2.97 |
| Bell | 3.43 | 0.84 - 6.02 | 1.31 | 0.0097 | 2.61 |
| CCC | -0.80 | -1.58 - -0.01 | 0.40 | 0.0467 | -1.99 |
| SF36 |  |  |  |  |  |
| Vitality | 3.29 | 0.53 - 6.05 | 1.40 | 0.0196 | 2.35 |
| General health | 3.41 | 0.81 - 6.00 | 1.32 | 0.0104 | 2.58 |

CI: Confidence interval. SD: Standard deviation. Only scores showing a significant treatment of BC007 are shown. The unit of the treatment effect is the unit of the corresponding questionnaire.

Means (SD, CI) of each score can be seen in supplementary tables 9-27. BC007 showed a significant treatment effect on self-reported fatigue (*FACIT Fatigue Scale*) until day 28 (Table 2). In addition, a significant improvement of the severity of fatigue was observed after BC007 treatment (*Bell score*, *Fatigue Severity Scale FSS*) until day 28 (Table 2). Scores for each visit can be seen in Figure 3. Considering inter-patients’ differences, crossover analysis showed a significant treatment effect of BC007 on fatigue (*FACIT Fatigue Scale*), the severity of fatigue (*Bell score*), and on ME/CFS diagnosis (*CCC*).

**Figure 3:**
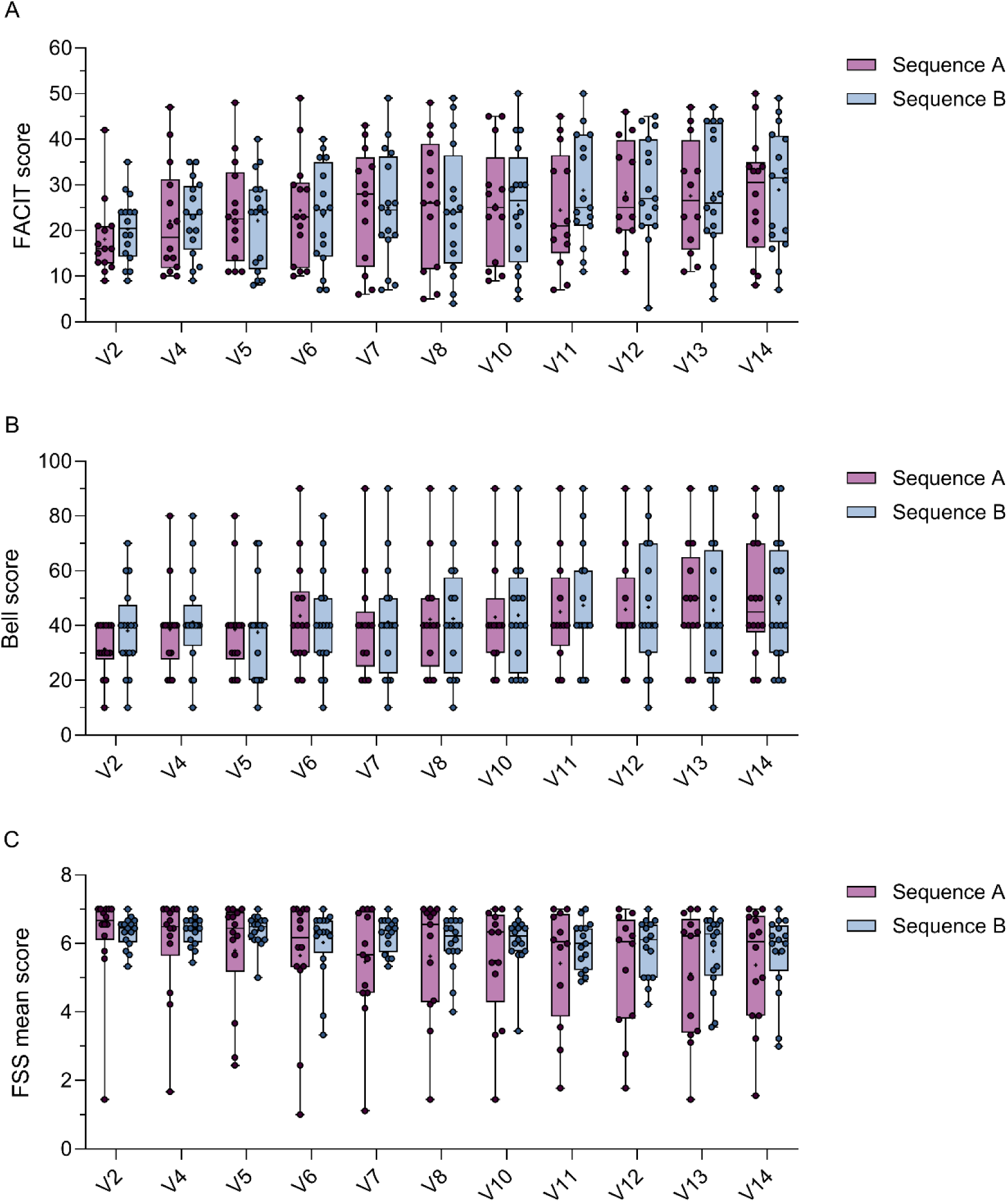
Fatigue scores assessed at V2, V4, V5, V6, V7, V8, V10, V11, V12, V13 and V14. (A) FACIT scores, (B) Bell scores and (C) FSS mean scores of study participants. Depicted are medians with IQR and ranges. Means are shown as crosses. Sequence A n V2-V6=14, n V7-V10=13, n V11-V12=12, n V13=13, n V14=14. Sequence B n V2-10=16, n V11-12=15, n V13-14=16. One patient in sequence A only received the first infusion (BC007) at V2 (ER28). One patient in sequence B only received the first infusion (placebo) at V2 (ER36).

Treatment effects of BC007 on quality of life showed a significantly improved vitality (*SF36 vitality*), general health (*SF36 general health*), and social role functioning (*SF36 social role function*) until day 28 (Table 2). A significant treatment effect of BC007 was observed on mental component summary (*SF36 MCS*) until day 28 (Table 2). Considering inter-patients’ differences, crossover analysis showed a significant treatment effect of BC007 on vitality (*SF36 vitality*) and general health (*SF36 general health,* Table 2). Scores for each visit can be seen in Figure 4 and suppl. tables 10-28.

**Figure 4:**
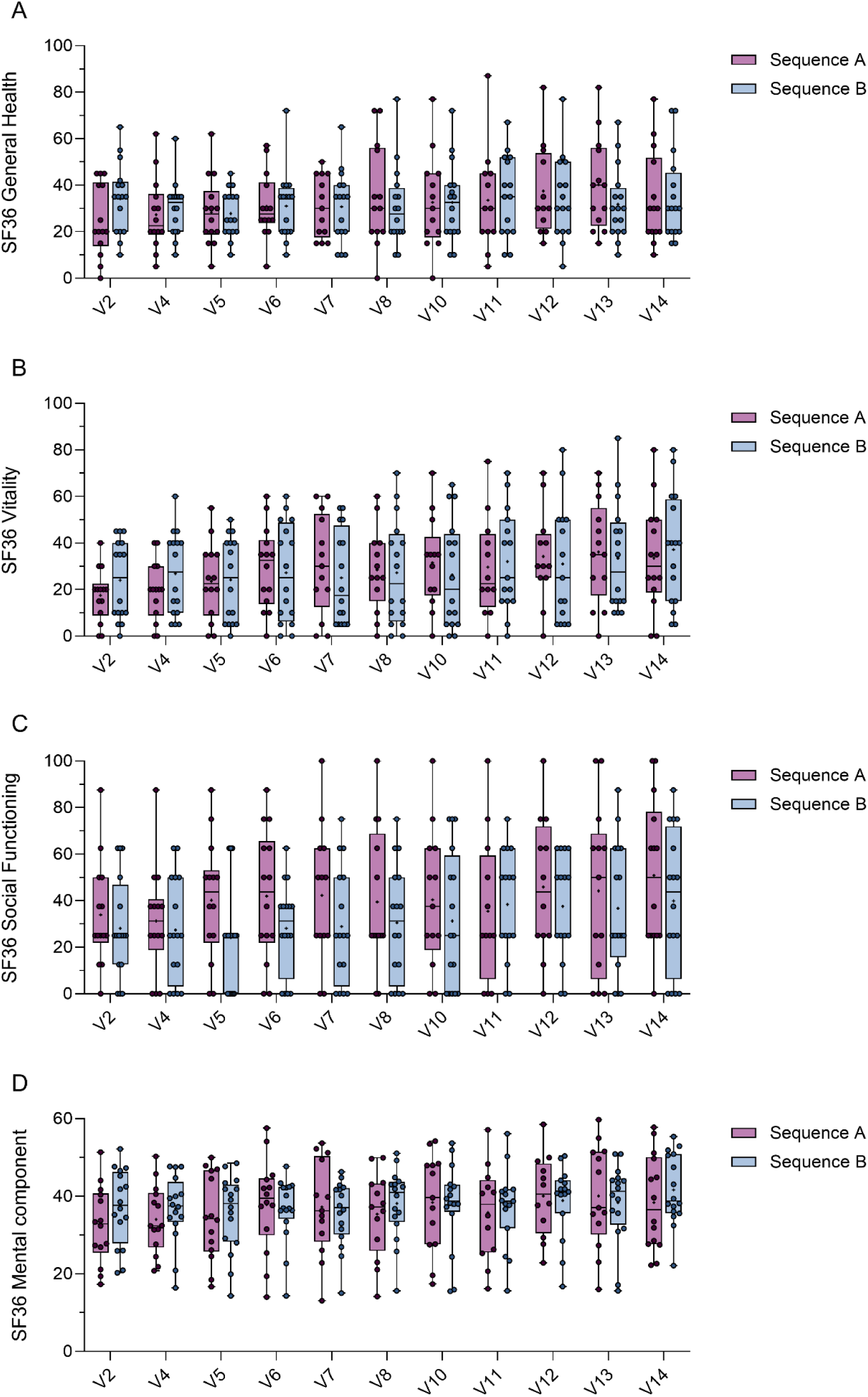
Selected SF36 scores assessed at V2, V4, V5, V6, V7, V8, V10, V11, V12, V13 and V14. (A) SF36 General health scores, (B) SF36 vitality scores, (C) SF36 social functioning scores and (D) SF36 mental component summary scores of study participants. Depicted are medians with IQR and ranges. Means are shown as crosses. Sequence A n V2-V6=14, n V7-V10=13, n V11-V12=12, n V13=13, n V14=14. Sequence B n V2-10=16, n V11-12=15, n V13-14=16. One patient in sequence A only received the first infusion (BC007) at V2 (ER28). One patient in sequence B only received the first infusion (placebo) at V2 (ER36).

No treatment effects of BC007 were observed on DSQ-PEM, walking distance (6MWT) and the subjective rating of level of exertion and dyspnoea during 6MWT (Borg scales). Individual patterns of GPCR-fAAb can be seen in Suppl. Table 29 and 30. Seropositivity of GPCR-fAAb was 100% (positive chronotropic: 100% β2-fAAb, 7% AT1-fAAb, 0% α1-fAAb, 0 % Noci-fAAb) and 100% (negative chronotropic: 0% MAS-AAb, 100% M2-AAb, 0% ETA-AAb) at screening (V1). In the 30 patients, both a quick (n=13, 43·3%) and delayed (n=7, 23·3%) effect of BC007 on GPCR-fAAb were observed. A third participants’ group (n=9, 30%) showed a seronegativity of GPCR-fAAb at d7 (sequence B), differing from the other participants in sequence B (Figure 5). One participant (3·3%) had a persistency of GPCR-fAAb within the study period (receiving only first infusion in sequence B). A relapse of GPCR-fAAB (d70) was observed in one participant (3·3%).

**Figure 5:**
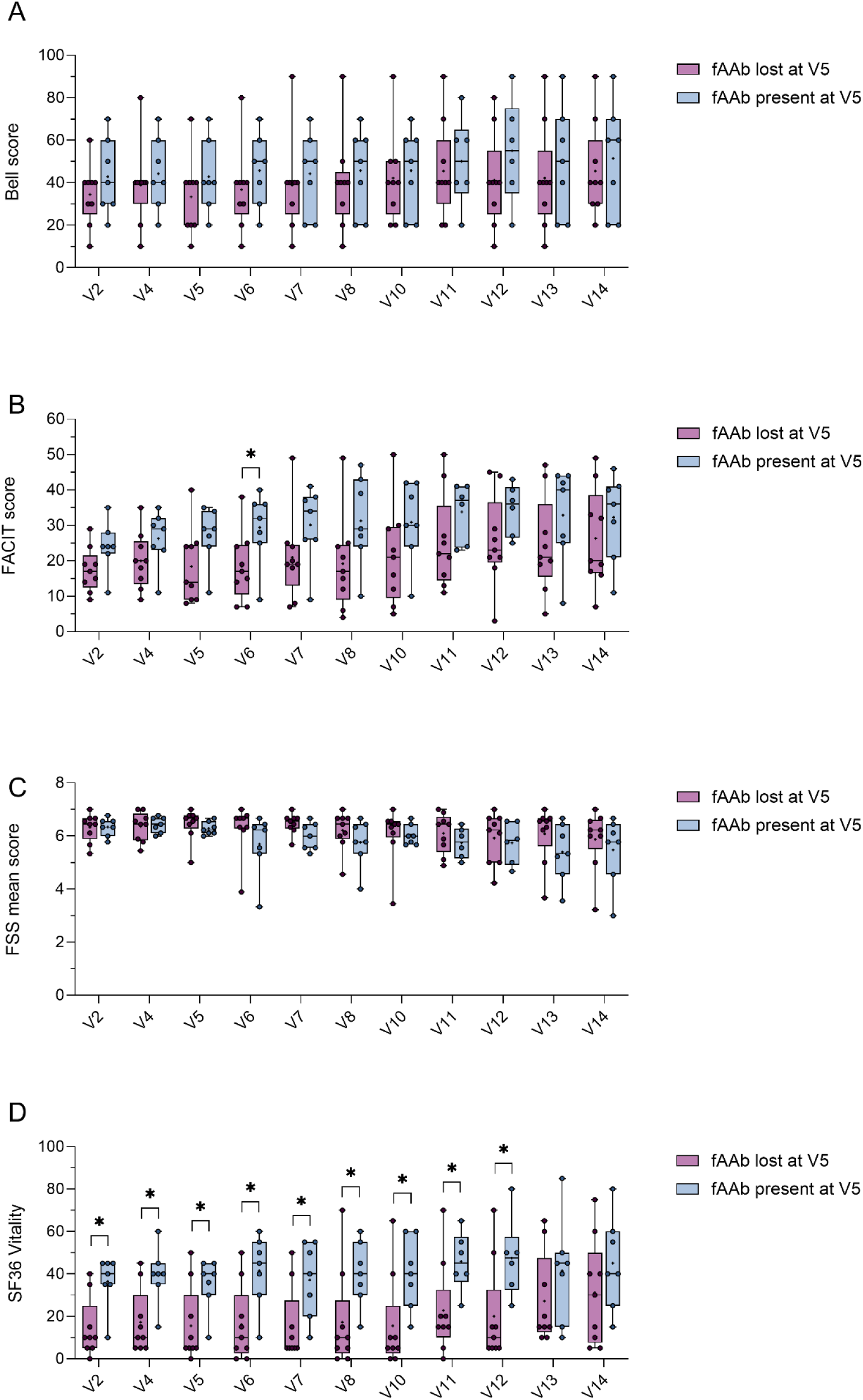
Selected Fatigue scores of study participants in sequence B, divided by fAAb status at Visit 5. n fAAb lost at V5=9, n fAAb present at V5 for V2-V10=7, n fAAb present at V5 for V11-12=6, n fAAb present at V5 for V13-14=7. One patient in sequence B only received the first infusion (placebo) at V2 (ER36). Mann-Whitney tests were used to determine statistical significance between the two groups. *=p≤0.05

## Discussion

The reCOVer study was an investigator-initiated, randomised, placebo-controlled clinical phase IIa trial to investigate safety, tolerability and clinical effects of BC007 on fatigue, severity of fatigue and quality of life in an autoimmune subgroup of PCS patients with GPCR-fAAbs. BC007 has been shown to neutralise GPCR-fAAbs, both in-vitro and in-vivo^18^ ^19^. Data published by us and others suggest an involvement of GPCR-fAAbs in the pathogenesis of PCS, leading to the hypothesis that neutralisation of such GPCR-fAAbs would improve PCS symptoms ^20^. The results of this clinical trial show a significant improvement of several fatigue scores after infusion of BC007, supporting this hypothesis. This is in line with an individual treatment of a PCS patient performed by our group, which also showed neutralisation of GPCR-fAAbs and improvement of PCS symptoms after BC007 infusion ^21^.

BC007 was well tolerated. Except for slight, clinically irrelevant, transient changes in blood anticoagulation, no relevant alteration of the laboratory parameters and no higher rate of AEs was observed compared to placebo, being in line with previous studies ^10^ ^22 23^. Notably, while we saw a significant improvement in fatigue assessed by the FACIT fatigue scale, Bell scale, FSS, and CCC; this was not the case with the Chalder fatigue scale. This data demonstrates clear differences between the different fatigue questionnaires.

While we observed a positive treatment effect of BC007, a Phase II clinical trial by Berlin Cures GmbH (BLOC) did not show a superior effect compared to placebo, as announced in a press release ^24^. Detailed participant characteristics and outcomes from BLOC have not been shared. Both trials included PCS patients with detectable GPCR-fAAbs, but there were key differences: BLOC used a randomised design, while reCOVer was a crossover study; inclusion criteria for fatigue severity differed (reCOVer: Bell Score≤60; BLOC: FACIT-FS Score<35); the duration of PCS symptoms varied (reCOVer: at least three months before enrolment; BLOC: „acute phase of COVID-19 ended at least 3 months prior to dosing“). Additionally, the reCOVer trial excluded patients with ‘organ damage.’ These differences may explain the discrepant results, though an in-depth analysis of the BLOC trial is needed to confirm this. No data on treatment success or detailed analysis have been provided from BLOC.

Current treatment trends emphasise a multidisciplinary approach, involving symptomatic, supportive, and rehabilitative care, with a significant role for patient self-monitoring and active participation in their recovery. Different clinical trials with PCS patients are running, differing in study protocols, patients’ characteristics and outcome variables (e. g. Fluvoxamine, Nirmatrelvir, (Methyl)prednisolone, Vortioxetine, Ivabradine, Naltrexone, Vericiguat)^25^. As an example, LTY-100 and Treamid are considered for patients with PCS with pulmonary fibrosis (i.e. subgroup organ damage). Treatment effects of Rintatolimod were considered for a subgroup of patients with PCS of severe PEM (ME/CFS).

The rationale for the treatment with BC007, an inhibitor of GPCR-fAAbs, was the assumption that GPCR-fAAbs exert a pathogenic effect in a subgroup of PCS patients. The continuous binding of GPCR-fAAb to their receptors not only keeps the receptors in a state of constant stimulation due to their agonistic effect, yet this continuous stimulation also leads to a decrease in the number of receptors on the cell surface and a further to a decreasing in mRNA levels ^26^, consequently affecting endothelial cell and circulating mononuclear cell function, muscles or nerves ^27^. Although patients received only a single infusion of BC007, the patients experienced a prolonged improvement of their symptoms. As BC007 has only a short serum half-life, we hypothesise that the drug not only prevents binding of the GPCR-fAAbs to their receptors, but in addition, it likely exerts an immunomodulating effect on B-cells and/or plasma cells. Further research is needed to delineate the assumed interaction of BC007 with antibody producing cells. Of interest a further participants’ subgroup showed a disappearance of GPCR-fAAb without BC007 treatment, yet not showing an improvement of fatigue in the following weeks, compared to an improvement of fatigue in those participants, whose GPCR-fAAb were neutralised after BC007. We assume that low serum levels of highly active GPCR-fAAb still have a functional pathogenic effect, even when their concentrations drop below the detection limit of the bioassay.

In contrast to BC007, immune adsorption (IA) non-specifically removes all immunoglobulins from patients’ plasma, including GPCR-fAAbs. Interim results of an observational study of repeated IA in patients with PCS of the subgroup ME/CFS showed a decrease in β2-fAAbs and an improvement for seven out of 10 patients as measured by the SF36-PF questionnaire ^28^. However, no control group was included. Currently, a double-blinded, randomised, sham-controlled IA study, including patients with ME/CFS, also after COVID-19, is being conducted though no results have been published yet ^29^. Disadvantages of IA, compared to BC007, are potential side effects due to the necessary puncture of large veins and the risk of infection due to the transient strong decrease of the plasma immunoglobulin levels which need several weeks to recover. In addition, repeated sessions in one week’s time makes IA for the patient more invasive than a one-time infusion of BC007.

ReCOVer is the first crossover study of BC007 in an autoimmune subgroup of patients with PCS with encouraging results. The crossover design enabled analysis of treatment effects in a low number of patients, additionally considering inter-patients’ differences in validated self-reported patients’ outcomes. The clinical trial was not designed to investigate effectiveness of BC007. Although the study is limited due to the low patient number and a short follow up duration of maximum 91 days, the present results of reCOVer encourage powering a subsequent clinical trial for investigation of effectiveness of BC007 and its long-term effects. An Intention-to-treat analysis (ITT) was performed including all randomised patients, with an underestimation of the treatment effect of BC007 due to inclusion of one participant of sequence B, who never received BC007 (so the treatment effects are conservative estimates).

Due to the heterogeneity of PCS, a uniform therapeutic procedure for all patients will most likely not be feasible. Thus, it is crucial to stratify patients into subgroups, based on objective biomarkers, in order to enable an effective PCS treatment. Some patients may benefit from symptomatic treatment and rehabilitation, while others with immune, autonomic, or capillary impairments will require different therapies. Stratification is the key to tailoring treatment for each patient.

## Conclusion

In conclusion, clinical trial reCOVer showed that BC007 exhibited an expected anticoagulatory effect directly after treatment. BC007 was well tolerated without serious adverse events in patients with PCS and showed a significant improvement of fatigue, severity of fatigue and quality of life. The data of the present clinical trial suggest that BC007 might offer a novel curative and causal therapy for an autoimmune subgroup of patients with PCS.

## Data Availability

All data produced in the present study are available upon reasonable request to the authors.

## Author contributions

Conceptualisation: BH, CM, TH, SM, MH, MG, JR

Methodology: BH, CM, SM, PL

Investigation: TH, JR, ASt, TK, VZ, ASk, MW, CM, SH, JS, MG

Formal analysis: AStr, KGS

Data curation: BH, AStr, MG, KGS

Visualisation: AStr, KGS, PL

Validation: BH, AStr, PL, KGS

Software: ER

Resources: BH, ASt, TK, ASk, VZ, TB, RHB, AB

Project administration: BH, PL

Funding acquisition: BH, CM, MG

Supervision: BH, CM, FK

Writing – original draft: BH, PL, KGS, CM, TH, JR, KGS, PL, MH

Writing – review & editing: All authors

## Acknowledgement

We thank Marie Albrecht, Barbara Meerwald, Melanie Pflügner, Lea Hack, Birgit Gerlach, Johannes Eissing, and Franziska Männel for their strong and continuous support within topics regarding the BC007 study trial.

## Notes

### Competing Interest Statement

The authors have declared no competing interest.

### Clinical Trial

EudraCT No.: 2022-001781-35

### Funding Statement

This study was funded by the German Federal Ministry of Education and Research, BMBF, grant number 01EP2108A (reCOVer) and 01EO2105 (iIMMUNE_ACS) as well as the German Research Foundation (DFG), grant number 401821119/GRK2504

### Author Declarations

Ethics committee of the Friedrich-Alexander-Universitaet Erlangen-Nuernberg, Erlangen, Germany gave ethical approval for this work.

## References

1. Soriano JB, Murthy S, Marshall JC, Relan P, Diaz JV, Definition WCC. A clinical case definition of post-COVID-19 condition by a Delphi consensus. Lancet Infect Dis 2022; 22(4): E102–E7.

2. Parotto M, Gyongyosi M, Howe K, et al. Post-acute sequelae of COVID-19: understanding and addressing the burden of multisystem manifestations. Lancet Respir Med 2023; 11(8): 739–54.

3. Proal AD, VanElzakker MB. Long COVID or Post-acute Sequelae of COVID-19 (PASC): An Overview of Biological Factors That May Contribute to Persistent Symptoms. Front Microbiol 2021; 12: 698169.

4. Kell DB, Laubscher GJ, Pretorius E. A central role for amyloid fibrin microclots in long COVID/PASC: origins and therapeutic implications. Biochem J 2022; 479(4): 537–59.

5. Ackermann M, Anders HJ, Bilyy R, et al. Patients with COVID-19: in the dark-NETs of neutrophils. Cell Death Differ 2021; 28(11): 3125–39.

6. Castanares-Zapatero D, Chalon P, Kohn L, et al. Pathophysiology and mechanism of long COVID: a comprehensive review. Ann Med 2022; 54(1): 1473–87.

7. Szewczykowski C, Mardin C, Lucio M, et al. Long COVID: Association of Functional Autoantibodies against G-Protein-Coupled Receptors with an Impaired Retinal Microcirculation. Int J Mol Sci 2022; 23(13).

8. Bock LC, Griffin LC, Latham JA, Vermaas EH, Toole JJ. Selection of single-stranded DNA molecules that bind and inhibit human thrombin. Nature 1992; 355(6360): 564–6.

9. Haberland A, Wallukat G, Schimke I. Aptamer binding and neutralization of beta1-adrenoceptor autoantibodies: basics and a vision of its future in cardiomyopathy treatment. Trends Cardiovasc Med 2011; 21(6): 177–82.

10. Becker NP, Haberland A, Wenzel K, et al. A Three-Part, Randomised Study to Investigate the Safety, Tolerability, Pharmacokinetics and Mode of Action of BC 007, Neutraliser of Pathogenic Autoantibodies Against G-Protein Coupled Receptors in Healthy, Young and Elderly Subjects. Clin Drug Investig 2020; 40(5): 433–47.

11. Cocks K, Torgerson DJ. Sample size calculations for pilot randomized trials: a confidence interval approach. J Clin Epidemiol 2013; 66(2): 197–201.

12. Berger VW, Ivanova A, Knoll MD. Minimizing predictability while retaining balance through the use of less restrictive randomization procedures. Stat Med 2003; 22(19): 3017–28.

13. Hohberger B, Harrer T, Mardin C, et al. Case Report: Neutralization of Autoantibodies Targeting G-Protein-Coupled Receptors Improves Capillary Impairment and Fatigue Symptoms After COVID-19 Infection. Front Med (Lausanne) 2021; 8: 754667.

14. Newcombe RG. Interval estimation for the difference between independent proportions: comparison of eleven methods. Stat Med 1998; 17(8): 873–90.

15. Frison L, Pocock SJ. Repeated measures in clinical trials: analysis using mean summary statistics and its implications for design. Stat Med 1992; 11(13): 1685–704.

16. Vickers AJ, Altman DG. Statistics notes: Analysing controlled trials with baseline and follow up measurements. BMJ 2001; 323(7321): 1123–4.

17. Senn SS. Cross-over Trials in Clinical Research; 2002.

18. Haberland A, Holtzhauer M, Schlichtiger A, et al. Aptamer BC 007 - A broad spectrum neutralizer of pathogenic autoantibodies against G-protein-coupled receptors. Eur J Pharmacol 2016; 789: 37–45.

19. Haberland A, Wallukat G, Dahmen C, Kage A, Schimke I. Aptamer neutralization of beta1-adrenoceptor autoantibodies isolated from patients with cardiomyopathies. Circulation research 2011; 109(9): 986–92.

20. Cabral-Marques O, Moll G, Catar R, et al. Autoantibodies targeting G protein-coupled receptors: An evolving history in autoimmunity. Report of the 4th international symposium. Autoimmunity reviews 2023; 22(5): 103310.

21. Hohberger B, Harrer T, Mardin C, et al. Case Report: Neutralization of Autoantibodies Targeting G-Protein-Coupled Receptors Improves Capillary Impairment and Fatigue Symptoms After COVID-19 Infection. Frontiers in Medicine 2021; 8.

22. Griffin LC, Tidmarsh GF, Bock LC, Toole JJ, Leung LL. In vivo anticoagulant properties of a novel nucleotide-based thrombin inhibitor and demonstration of regional anticoagulation in extracorporeal circuits. Blood 1993; 81(12): 3271–6.

23. DeAnda A, Jr., Coutre SE, Moon MR, et al. Pilot study of the efficacy of a thrombin inhibitor for use during cardiopulmonary bypass. Ann Thorac Surg 1994; 58(2): 344–50.

24. Website of Berlin Cures. 2024. https://www.berlincures.com/en/news/phase-2-long-covid-results.

25. Yong SJ, Halim A, Halim M, et al. Experimental drugs in randomized controlled trials for long-COVID: what’s in the pipeline? A systematic and critical review. Expert Opin Investig Drugs 2023; 32(7): 655–67.

26. Podlowski S, Luther HP, Morwinski R, Muller J, Wallukat G. Agonistic anti-beta1-adrenergic receptor autoantibodies from cardiomyopathy patients reduce the beta1-adrenergic receptor expression in neonatal rat cardiomyocytes. Circulation 1998; 98(22): 2470–6.

27. Sotzny F, Filgueiras IS, Kedor C, et al. Dysregulated autoantibodies targeting vaso-and immunoregulatory receptors in Post COVID Syndrome correlate with symptom severity. Frontiers in immunology 2022; 13: 981532.

28. Stein E, Heindrich C, Wittke K, et al. Observational Study of Repeat Immunoadsorption (RIA) in Post-COVID ME/CFS Patients with Elevated ss2-Adrenergic Receptor Autoantibodies-An Interim Report. J Clin Med 2023; 12(19).

29. Pressler H, Machule ML, Ufer F, et al. IA-PACS-CFS: a double-blinded, randomized, sham-controlled, exploratory trial of immunoadsorption in patients with chronic fatigue syndrome (CFS) including patients with post-acute COVID-19 CFS (PACS-CFS). Trials 2024; 25(1): 172.

