## Supplementary Information for "Safety, tolerability and clinical effects of BC007 (Rovunaptabin) on fatigue and quality of life in patients with post-COVID syndrome (reCOVer): a prospective, exploratory, randomised, placebo-controlled, double-blind, crossover phase IIa clinical trial"

**Supplementary Figures**

Supplementary Figure 1: Coagulation status of study participants.

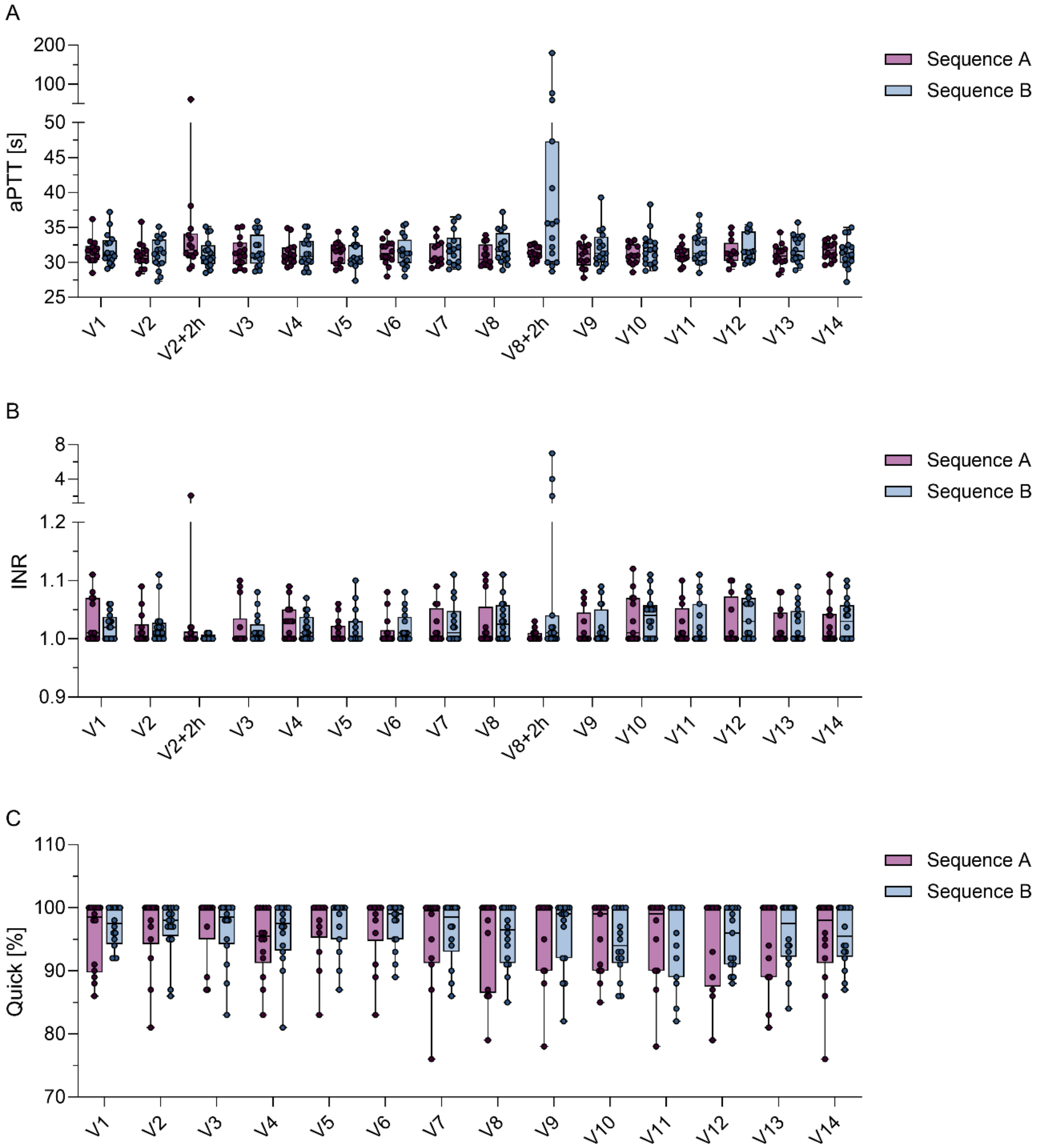


**Supplementary Figure 1:** Coagulation status of study participants. (A) aPTT values of study participants in seconds, measured at visits V1-V14 as well as 2h after the infusions at V2 and V8. (B) INR values of study participants, measured at visits V1-V14 as well as 2h after the infusions at V2 and V8. (A) Quick values of study participants in percentages, measured at visits V1-V14. n Sequence A: 14, n Sequence B: 16. aPTT: activated partial thromboplastin time, INR: International normalised ratio.

**Supplementary Tables**

Supplementary Table 1: Overview of comorbidities of study participants.

Supplementary Table 2: Comorbidities of study participants.

Supplementary Table 3: List of co-medications.

Supplementary Table 4: COVID-19 vaccinations of study participants.

Supplementary Table 5: aPTT values of study participants.

Supplementary Table 6: INR values of study participants.

Supplementary Table 7: Quick values of study participants.

Supplementary Table 8: Adverse events observed in study participants.

Supplementary Table 9: Descriptive list of adverse events.

Supplementary Table 10: FACIT scores of study participants.

Supplementary Table 11: Bell scores of study participants.

Supplementary Table 12: FSS scores of study participants.

Supplementary Table 13: Canadian consensus criteria (CCC) scores of study participants.

Supplementary Table 14: Vitality scores of study participants assessed by SF36 questionnaire.

Supplementary Table 15: General Health scores of study participants assessed by SF36 questionnaire.

Supplementary Table 16: Social role functioning scores of study participants assessed by SF36 questionnaire.

Supplementary Table 17: Mental component summary (MCS) scores of study participants assessed by SF36 questionnaire.

Supplementary Table 18: Physical function scores of study participants assessed by SF36 questionnaire.

Supplementary Table 19: Physical role function scores of study participants assessed by SF36 questionnaire.

Supplementary Table 20: Emotional role functioning scores of study participants assessed by SF36 questionnaire.

Supplementary Table 21: Emotional well-being scores of study participants assessed by SF36 questionnaire.

Supplementary Table 22: Physical pain scores of study participants assessed by SF36 questionnaire.

Supplementary Table 23: Physical component summary (PCS) scores of study participants assessed by SF36 questionnaire.

Supplementary Table 24: Chalder Fatigue scale scores of study participants.

Supplementary Table 25: Walking distance of study participants in meters assessed by 6MWT.

Supplementary Table 26: Exertion of study participants assessed by 6MWT.

Supplementary Table 27: Dyspnoea of study participants assessed by 6MWT.

Supplementary Table 28: DSQ-PEM scores of study participants.

Supplementary Table 29: Positive and negative chronotropic fAAbs of study participants.

Supplementary Table 30: GPCR-fAAbs of study participants.

**Supplementary Table 1:** Overview of comorbidities of study participants.

| | Sequence A | Sequence B | p-value | Effect size | Total |
|---|---|---|---|---|---|
| **Comorbidities** | | | | | |
| Yes | 42.9% ( 6) | 43.8% ( 7) | 1.000 | 0.97 (0.18, 5.13) | 43.3% (13) |
| No | 57.1% ( 8) | 56.2% ( 9) | | | 56.7% (17) |
| Missing | 0 | 0 | | | 0 |
| **Allergy** | | | | | |
| Yes | 21.4% ( 3) | 6.2% ( 1) | 0.495 | 3.91 (0.27, 228.44) | 13.3% ( 4) |
| No | 78.6% (11) | 93.8% (15) | | | 86.7% (26) |
| Missing | 0 | 0 | | | 0 |
| **Pollen allergy** | | | | | |
| Yes | 14.3% ( 2) | 12.5% ( 2) | 1.000 | 1.16 (0.07, 18.32) | 13.3% ( 4) |
| No | 85.7% (12) | 87.5% (14) | | | 86.7% (26) |
| Missing | 0 | 0 | | | 0 |
| **Food allergy** | | | | | |
| Yes | 7.1% ( 1) | 6.2% ( 1) | 1.000 | 1.15 (0.01, 96.46) | 6.7% ( 2) |
| No | 92.9% (13) | 93.8% (15) | | | 93.3% (28) |
| Missing | 0 | 0 | | | 0 |
| **Drug allergy** | | | | | |
| Yes | 14.3% ( 2) | 6.2% ( 1) | 0.903 | 2.43 (0.11, 156.98) | 10.0% ( 3) |
| No | 85.7% (12) | 93.8% (15) | | | 90.0% (27) |
| Missing | 0 | 0 | | | 0 |
| **Diabetes Mellitus** | | | | | |
| Yes | 0.0% ( 0) | 0.0% ( 0) | NaN | 0.00 (0.00, Inf) | 0.0% ( 0) |
| No | 100.0% (14) | 100.0% (16) | | | 100.0% (30) |
| Missing | 0 | 0 | | | 0 |
| **Pulmonology** | | | | | |
| Yes | 0.0% ( 0) | 12.5% ( 2) | 0.525 | 0.00 (0.00, 6.05) | 6.7% ( 2) |
| No | 100.0% (14) | 87.5% (14) | | | 93.3% (28) |
| Missing | 0 | 0 | | | 0 |
| **Rheumatology** | | | | | |
| Yes | 0.0% ( 0) | 0.0% ( 0) | NaN | 0.00 (0.00, Inf) | 0.0% ( 0) |
| No | 100.0% (14) | 100.0% (16) | | | 100.0% (30) |
| Missing | 0 | 0 | | | 0 |
| **Coronary heart disease** | | | | | |
| Yes | 0.0% ( 0) | 0.0% ( 0) | NaN | 0.00 (0.00, Inf) | 0.0% ( 0) |
| No | 100.0% (14) | 100.0% (16) | | | 100.0% (30) |
| Missing | 0 | 0 | | | 0 |
| **Arterial hypertension** | | | | | |
| Yes | 0.0% ( 0) | 12.5% ( 2) | 0.525 | 0.00 (0.00, 6.05) | 6.7% ( 2) |
| No | 100.0% (14) | 87.5% (14) | | | 93.3% (28) |
| Missing | 0 | 0 | | | 0 |
| **Cardiac arrhythmia** | | | | | |
| Yes | 7.1% ( 1) | 12.5% ( 2) | 1.000 | 0.55 (0.01, 11.75) | 10.0% ( 3) |
| No | 92.9% (13) | 87.5% (14) | | | 90.0% (27) |
| Missing | 0 | 0 | | | 0 |
| **Cardiac insufficiency** | | | | | |
| Yes | 0.0% ( 0) | 0.0% ( 0) | NaN | 0.00 (0.00, Inf) | 0.0% ( 0) |
| No | 100.0% (14) | 100.0% (16) | | | 100.0% (30) |
| Missing | 0 | 0 | | | 0 |
| **Other endocrinal diseases** | | | | | |
| Yes | 7.1% ( 1) | 12.5% ( 2) | 1.000 | 0.55 (0.01, 11.75) | 10.0% ( 3) |
| No | 92.9% (13) | 87.5% (14) | | | 90.0% (27) |
| Missing | 0 | 0 | | | 0 |
| **Haematological/oncological diseases** | | | | | |
| Yes | 0.0% ( 0) | 0.0% ( 0) | NaN | 0.00 (0.00, Inf) | 0.0% ( 0) |
| No | 100.0% (14) | 100.0% (16) | | | 100.0% (30) |
| Missing | 0 | 0 | | | 0 |
| **Neurological diseases** | | | | | |
| Yes | 7.1% ( 1) | 0.0% ( 0) | 0.946 | Inf (0.03, Inf) | 3.3% ( 1) |
| No | 92.9% (13) | 100.0% (16) | | | 96.7% (29) |

| | | | | | |
|---|---|---|---|---|---|
| Missing | 0 | 0 | | | 0 |
| **Urological diseases** | | | | | |
| Yes | 0.0% ( 0) | 12.5% ( 2) | 0.525 | 0.00 (0.00, 6.05) | 6.7% ( 2) |
| No | 100.0% (14) | 87.5% (14) | | | 93.3% (28) |
| Missing | 0 | 0 | | | 0 |
| **Infectious diseases** | | | | | |
| Yes | 21.4% ( 3) | 18.8% ( 3) | 1.000 | 1.18 (0.13, 10.67) | 20.0% ( 6) |
| No | 78.6% (11) | 81.2% (13) | | | 80.0% (24) |
| Missing | 0 | 0 | | | 0 |

NaN: Not a number, Inf: Infinite.

**Supplementary Table 2:** Comorbidities of study participants.

| | **Comorbidity** | **Co-medication** |
|---|---|---|
| **ER-01** | Post-COVID | No |
| | Tachycardia | Yes |
| **ER-02** | Post-COVID | Yes |
| | POTS | No |
| **ER-03** | Post-COVID | No |
| **ER-04** | Post-COVID | No |
| | Hypothyreosis | Yes |
| **ER-05** | Post-COVID | No |
| **ER-06** | Post-COVID | No |
| | Pancolitis ulcerosa | Yes |
| **ER-07** | Post-COVID | Yes |
| **ER-09** | Post-COVID | No |
| | Bankart lesion right shoulder | No |
| **ER-10** | Post-COVID | Yes |
| **ER-11** | Post-COVID | No |
| **ER-12** | Post-COVID | No |
| **ER-13** | Post-COVID | Yes |
| **ER-15** | Post-COVID | No |
| **ER-16** | Post-COVID | Yes |
| **ER-17** | Post-COVID | No |
| | Prostatic hyperplasia | Yes |
| | Irritable bowel syndrome | No |
| **ER-18** | Post-COVID | No |
| | Hypertension | Yes |
| **ER-20** | Post-COVID | No |
| | Neurodermatitis | No |
| **ER-21** | Post-COVID | No |
| **ER-23** | Post-COVID | No |
| | Bronchial asthma | No |
| | Arterial hypertension | Yes |
| | Chronic pain syndrome | No |
| **ER-24** | Post-COVID | No |
| **ER-27** | Post-COVID | No |
| **ER-28** | Post-COVID | No |
| **ER-29** | Post-COVID | No |
| | Hypothyreosis | Yes |
| **ER-31** | Post-COVID | Yes |
| | Arterial hypertension | Yes |
| **ER-32** | Post-COVID | Yes |
| **ER-33** | Post-COVID | No |
| **ER-34** | Post-COVID | No |
| | Herpes genitalis | Yes |
| **ER-35** | Post-COVID | Yes |
| **ER-36** | Post-COVID | No |
| | Histamine intolerance | No |
| **ER-37** | Post-COVID | No |
| | Hashimoto thyroiditis | Yes |
| | Endometriosis | No |

POTS: Postural orthostatic tachycardia syndrome, EBV: Epstein-Barr virus, N/A: None available

**Supplementary Table 3:** Co-medications of study participants.

| | Co-medication | Substance | Indication |
|---|---|---|---|
| **ER01** | Yes | Carvedilol | Tachycardia |
| **ER02** | Yes | Naltrexone<br>Aripiprazole | Post-COVID<br>Post-COVID |
| **ER03** | No | | |
| **ER04** | Yes | L-thyroxine<br>Diclofenac<br>Oseltamivir<br>Lidocaine | Hypothyreosis<br>AE Nr. 1 (Anal vein thrombosis)<br>AE Nr. 2 (Influenza A)<br>AE Nr. 1 (Anal vein thrombosis) |
| **ER05** | Yes | Ibuprofen | Prophylaxis |
| **ER06** | Yes | Carvedilol<br>Tofacitinib<br>Metamizole<br>Pregabalin<br>Mesalazine | Prophylaxis<br>Pancolitis ulcerosa<br>Prophylaxis<br>Prophylaxis<br>Pancolitis ulcerosa |
| **ER07** | Yes | Lidocaine<br>Ivabradine<br>Aripiprazole | Post-COVID<br>Post-COVID<br>Post-COVID |
| **ER09** | Yes | Ciprofloxacin | AE Nr. 1 (Epidymorchitis) |
| **ER10** | Yes | Beclometason/Formoterol spray<br>Losartan<br>Paracetamol | Post-COVID<br>Post-COVID<br>AE Nr. 2 (Cold) |
| **ER11** | Yes | Paracetamol | AE Nr. 3 (Gastrointestinal infection) |
| **ER12** | No | | |
| **ER13** | Yes | Low dose naltrexone | Post-COVID |
| **ER15** | No | | |
| **ER16** | Yes | Naltrexone | Post-COVID |
| **ER17** | Yes | Granufink Prosta forte | Prostatic hyperplasia |
| **ER18** | Yes | Enoxaparin<br>Candesartan<br>Heparin ointment | AE Nr. 1 (Thrombophlebitis)<br>Hypertension<br>AE Nr. 1 (Thrombophlebitis) |
| **ER20** | No | | |
| **ER21** | No | | |
| **ER23** | Yes | Candesartan | Arterial hypertension |
| **ER24** | No | | |
| **ER27** | No | | |
| **ER28** | No | | |
| **ER29** | Yes | L-thyroxine | Hypothyreose |
| **ER31** | Yes | Candesartan<br>Serrapeptase | Arterial hypertension<br>Post-COVID |
| **ER32** | Yes | Naltrexone | Prophylaxis |
| **ER33** | Yes | Cetirizine<br>Adrenaline<br>Betamethasone<br>Cortisone | AE Nr. 1 (Wasp sting)<br>AE Nr. 1 (Wasp sting)<br>AE Nr. 1 (Wasp sting)<br>AE Nr. 1 (Wasp sting) |
| **ER34** | Yes | Valaciclovir | Herpes genitalis |
| **ER35** | Yes | Simvastatin | Post-COVID |
| **ER36** | Yes | Clemastine<br>Prednisolone<br>Famotidine<br>Loratadine | AE Nr. 1,2 (Numbness of lips, face and hand, tightness of throat)<br>AE Nr. 1,2 (Numbness of lips, face and hand, tightness of throat)<br>AE Nr. 1,2 (Numbness of lips, face and hand, tightness of throat)<br>AE Nr. 1,2 (Numbness of lips, face and hand, tightness of throat) |
| **ER37** | Yes | L-thyroxin<br>Low dose naltrexone | Hashimoto thyroiditis<br>Prophylaxis |

ER28 only received one infusion (BC007) at V2 (d0). ER36 only received one infusion (placebo) at V2 (d0).

**Supplementary Table 4:** COVID-19 vaccinations of study participants.

| | Sequence A | Sequence B | p-value | Effect size | Total |
|---|---|---|---|---|---|
| **Number of vaccinations** | | | | | |
| 0 | 14% ( 2) | 6% ( 1) | | | 10% ( 3) |
| 1 | 7% ( 1) | 0% ( 0) | | | 3% ( 1) |
| 2 | 43% ( 6) | 56% ( 9) | 0.619 | 0.30 (0.00, 0.60) | 50% (15) |
| 3 | 21% ( 3) | 31% ( 5) | | | 27% ( 8) |
| 4 | 14% ( 2) | 6% ( 1) | | | 10% ( 3) |
| Missing | 0 | 0 | | | 0 |
| **Type of vaccine at 1st dose** | | | | | |
| BNT162b2 mRNA | 91.7% (11) | 86.7% (13) | | | 88.9% (24) |
| mRNA-1273 | 8.3% ( 1) | 6.7% ( 1) | | | 7.4% ( 2) |
| ChAdOx1-S | 0.0% ( 0) | 6.7% ( 1) | | | 3.7% ( 1) |
| Gam-COVID-Vac | 0.0% ( 0) | 0.0% ( 0) | 0.656 | NaN (NaN, NaN) | 0.0% ( 0) |
| Ad26.COV2.S | 0.0% ( 0) | 0.0% ( 0) | | | 0.0% ( 0) |
| No vaccination | 0.0% ( 0) | 0.0% ( 0) | | | 0.0% ( 0) |
| Missing | 2 | 1 | | | 3 |
| **Type of vaccine at 2nd dose** | | | | | |
| BNT162b2 mRNA | 81.8% ( 9) | 80.0% (12) | | | 80.8% (21) |
| mRNA-1273 | 18.2% ( 2) | 20.0% ( 3) | | | 19.2% ( 5) |
| ChAdOx1-S | 0.0% ( 0) | 0.0% ( 0) | | | 0.0% ( 0) |
| Gam-COVID-Vac | 0.0% ( 0) | 0.0% ( 0) | 1.000 | NaN (NaN, NaN) | 0.0% ( 0) |
| Ad26.COV2.S | 0.0% ( 0) | 0.0% ( 0) | | | 0.0% ( 0) |
| No vaccination | 0.0% ( 0) | 0.0% ( 0) | | | 0.0% ( 0) |
| Missing | 3 | 1 | | | 4 |
| **Type of vaccine at 3rd dose** | | | | | |
| BNT162b2 mRNA | 80.0% ( 4) | 33.3% ( 2) | | | 54.5% ( 6) |
| mRNA-1273 | 20.0% ( 1) | 66.7% ( 4) | | | 45.5% ( 5) |
| ChAdOx1-S | 0.0% ( 0) | 0.0% ( 0) | | | 0.0% ( 0) |
| Gam-COVID-Vac | 0.0% ( 0) | 0.0% ( 0) | 0.347 | NaN (NaN, NaN) | 0.0% ( 0) |
| Ad26.COV2.S | 0.0% ( 0) | 0.0% ( 0) | | | 0.0% ( 0) |
| No vaccination | 0.0% ( 0) | 0.0% ( 0) | | | 0.0% ( 0) |
| Missing | 9 | 10 | | | 19 |
| **Type of vaccine at 4th dose** | | | | | |
| BNT162b2 mRNA | 100.0% ( 2) | 100.0% ( 1) | | | 100.0% ( 3) |
| mRNA-1273 | 0.0% ( 0) | 0.0% ( 0) | | | 0.0% ( 0) |
| ChAdOx1-S | 0.0% ( 0) | 0.0% ( 0) | | | 0.0% ( 0) |
| Gam-COVID-Vac | 0.0% ( 0) | 0.0% ( 0) | NaN | NaN (NaN, NaN) | 0.0% ( 0) |
| Ad26.COV2.S | 0.0% ( 0) | 0.0% ( 0) | | | 0.0% ( 0) |
| No vaccination | 0.0% ( 0) | 0.0% ( 0) | | | 0.0% ( 0) |
| Missing | 12 | 15 | | | 27 |

NaN: Not a number.

**Supplementary Table 5:** aPTT values of study participants.

| | | Sequence A | | Sequence B | |
|---|---|---|---|---|---|
| | n | Mean | SD | Mean | SD |
| V2[1] | 14 | 31.02 | 1.86 | 31.24 | 2.25 |
| V2 + 2h[2] | 14 | 34.29 | 8.081 | 31.28 | 2.25 |
| V3 | 14 | 31.40 | 2.07 | 31.73 | 2.29 |
| V4 | 14 | 31.34 | 1.71 | 31.47 | 2.14 |
| V5 | 14 | 31.42 | 1.54 | 30.93 | 1.84 |
| V6 | 14 | 31.21 | 1.73 | 31.49 | 2.20 |
| V7 | 12 | 31.58 | 1.72 | 31.96 | 2.41 |
| V8[1] | 13 | 30.95 | 1.61 | 32.11 | 2.30 |
| V8 + 2h[2] | 13 | 31.45 | 0.95 | 48.38 | 38.71 |
| V9 | 13 | 30.95 | 1.72 | 32.01 | 2.67 |
| V10 | 13 | 31.04 | 1.44 | 31.82 | 2.44 |
| V11 | 12 | 31.19 | 1.37 | 32.15 | 2.31 |
| V12 | 12 | 31.61 | 1.74 | 31.90 | 1.94 |
| V13 | 13 | 30.98 | 1.60 | 31.93 | 1.93 |
| V14 | 13 | 31.62 | 1.35 | 31.38 | 2.07 |

[1]Blood was drawn before infusion of BC007 or placebo. [2]Blood was drawn two hours after infusion of BC007 or placebo. aPTT: activated partial thromboplastin time, given in seconds. SD: Standard deviation.

**Supplementary Table 6:** INR values of study participants.

| | | Sequence A | | Sequence B | |
|---|---|---|---|---|---|
| | n | Mean | SD | Mean | SD |
| V2[1] | 14 | 1.03 | 0.05 | 1.02 | 0.03 |
| V2 + 2h[2] | 14 | 1.08 | 0.29 | 1.00 | 0.01 |
| V3 | 14 | 1.02 | 0.04 | 1.02 | 0.04 |
| V4 | 14 | 1.04 | 0.04 | 1.03 | 0.04 |
| V5 | 14 | 1.02 | 0.04 | 1.02 | 0.03 |
| V6 | 14 | 1.02 | 0.04 | 1.02 | 0.03 |
| V7 | 12 | 1.04 | 0.06 | 1.03 | 0.04 |
| V8[1] | 13 | 1.04 | 0.06 | 1.03 | 0.03 |
| V8 + 2h[2] | 13 | 1.01 | 0.01 | 1.68 | 1.68 |
| V9 | 13 | 1.03 | 0.06 | 1.03 | 0.04 |
| V10 | 13 | 1.03 | 0.04 | 1.04 | 0.04 |
| V11 | 12 | 1.04 | 0.06 | 1.04 | 0.05 |
| V12 | 12 | 1.04 | 0.06 | 1.04 | 0.04 |
| V13 | 13 | 1.04 | 0.05 | 1.03 | 0.04 |
| V14 | 13 | 1.04 | 0.06 | 1.03 | 0.03 |

[1]Blood was drawn before infusion of BC007 or placebo. [2]Blood was drawn two hours after infusion of BC007 or placebo. INR: International normalized ratio. SD: Standard deviation.

**Supplementary Table 7:** Quick values of study participants.

| | | Sequence A | | Sequence B | |
|---|---|---|---|---|---|
| | n | Mean | SD | Mean | SD |
| V2[1] | 14 | 96.43 | 5.91 | 96.75 | 4.34 |
| V3 | 14 | 97.14 | 5.22 | 96.50 | 5.11 |
| V4 | 14 | 94.71 | 5.43 | 96.25 | 5.26 |
| V5 | 14 | 96.93 | 5.08 | 97.20 | 4.18 |
| V6 | 14 | 96.86 | 5.36 | 97.31 | 3.61 |
| V7 | 12 | 95.00 | 7.48 | 96.21 | 4.95 |
| V8[1] | 13 | 94.77 | 7.45 | 95.50 | 4.75 |
| V9 | 13 | 95.46 | 6.98 | 95.88 | 5.68 |
| V10 | 13 | 95.23 | 5.63 | 94.31 | 5.00 |
| V11 | 12 | 94.83 | 7.15 | 95.00 | 6.47 |
| V12 | 12 | 94.58 | 7.45 | 95.00 | 4.80 |
| V13 | 13 | 94.38 | 7.04 | 95.69 | 5.12 |
| V14 | 13 | 94.92 | 7.43 | 95.50 | 4.70 |

[1]Blood was drawn before infusion of BC007 or placebo. Values are given in percentages. SD: Standard deviation.

**Supplementary Table 8:** Adverse events observed in study participants.

| Patient | Sequence | AE | Day of AE | Severity | Reasonable causal relation | Serious AE | System/Organ class | Preferred Term |
|---|---|---|---|---|---|---|---|---|
| ER-02 | A | Increased GOT | 1 | Mild | No | No | Investigations | Aspartate aminotransferase increased |
| ER-02 | A | Increased GPT | 1 | Mild | No | No | Investigations | Alanine aminotransferase increased |
| ER-02 | A | Seasonal Infection / Cold | 64 | Mild | No | No | Infections and infestations | Nasopharyngitis |
| ER-02 | A | Periodontitis | 64 | Mild | No | No | Infections and infestations | Periodontitis |
| ER-04 | B | Anal vein thrombosis | 20 | Mild | No | No | Gastrointestinal disorders | Haemorrhoids thrombosed |
| ER-04 | B | Influenza A | 28 | Moderate | No | No | Infections and infestations | Influenza |
| ER-06 | A | Mild infection | 20 | Mild | No | No | Infections and infestations | Nasopharyngitis |
| ER-06 | A | Deterioration of Colitis Ulcerosa | 24 | Mild | No | No | Gastrointestinal disorders | Colitis ulcerative |
| ER-07 | B | Impaired coagulation | 42 | Moderate | Yes | No | Investigations | Coagulation test abnormal |
| ER-09 | B | Epidymorchitis | 42 | Mild | No | No | Infections and infestations | Orchitis |
| ER-10 | A | Cold | 16 | Moderate | No | No | Infections and infestations | Nasopharyngitis |
| ER-10 | A | Cold | 77 | Mild | No | No | Infections and infestations | Nasopharyngitis |
| ER-11 | A | Increased INR | 0 | Mild | Yes | No | Investigations | International normalised ratio increased |
| ER-11 | A | Increased aPTT | 0 | Mild | Yes | No | Investigations | Activated partial thromboplastin time prolonged |
| ER-11 | A | Gastrointestinal infection | 42 | Mild | No | No | Infections and infestations | Gastrointestinal infection |
| ER-12 | B | Palpable Lymph Node | 88 | Mild | No | No | Investigations | Lymph node palpable |
| ER-13 | B | Impaired INR | 43 | Mild | Yes | No | Investigations | International normalised ratio abnormal |
| ER-15 | B | Impaired INR | 42 | Mild | Yes | No | Investigations | International normalised ratio abnormal |
| ER-17 | A | Angina Pectoris | 44 | Mild | No | No | Cardiac disorders | Angina pectoris |
| ER-17 | A | Gastric pain | 48 | Mild | No | No | Gastrointestinal disorders | Abdominal pain upper |
| ER-18 | B | Thrombophlebitis | 43 | Mild | No | No | Vascular disorders | Thrombophlebitis |
| ER-21 | B | Dyspnoe | 41 | Moderate | No | No | Respiratory, thoracic and mediastinal disorders | Dyspnoea |
| ER-28 | A | Flu-like Infect | 25 | Moderate | No | No | General disorders and administration site conditions | Influenza like illness |
| ER-28 | A | Corona Re-Infection | 38 | Moderate | No | No | Infections and infestations | COVID-19 |
| ER-33 | A | Wasp sting | 33 | Mild[1] | No | Yes[1] | Injury, poisoning and procedural complications | Arthropod sting |
| ER-33 | A | Common Cold | 42 | Mild | No | No | Infections and infestations | Nasopharyngitis |
| ER-34 | A | Depressive mood | 0 | Mild | No | No | Psychiatric disorders | Depressed mood |
| ER-34 | A | Flu-like Infect | 24 | Mild | No | No | General disorders and administration site conditions | Influenza like illness |
| ER-36 | B | Numbness of lips, face and hand | 0 | Moderate | Yes | No | Nervous system disorders | Hypoaesthesia |

| | | | | | | | | |
|---|---|---|---|---|---|---|---|---|
| ER-36 | B | Tightness of throat | 0 | Moderate | Yes | No | Respiratory, thoracic and mediastinal disorders | Throat tightness |
| ER-37 | B | Higher Lipase Values in Bloodwork | 14 | Mild | No | No | Investigations | Lipase increased |

[1]Patient had mild symptoms but stayed overnight in the hospital for monitoring due to a known wasp venom allergy, thus the AE was categorized as serious. AE: Adverse event, INR: International normalised ratio, aPTT: activated partial thromboplastin time. ER28 only received one infusion (BC007) at V2 (d0). ER36 only received one infusion (placebo) at V2 (d0).

**Supplementary Table 9:** Descriptive list of adverse events.

| | Sequence A | Sequence B | p-value | Effect size | Total 31 AEs |
|---|---|---|---|---|---|
| **Severity** | | | | | |
| Mild | 84% (16) | 58% ( 7) | 0.237 | NaN ( NaN, NaN) | 74% (23) |
| Moderate | 16% ( 3) | 42% ( 5) | | | 26% ( 8) |
| Severe | 0% ( 0) | 0% ( 0) | | | 0% ( 0) |
| Life-threatening | 0% ( 0) | 0% ( 0) | | | 0% ( 0) |
| Death | 0% ( 0) | 0% ( 0) | | | 0% ( 0) |
| Missing | 0 | 0 | | | 0 |
| **Causal relation** | | | | | |
| Reasonable possibility | 11% ( 2) | 42% ( 5) | 0.114 | 0.18 (0.01, 1.38) | 23% ( 7) |
| No reasonable possibility | 89% (17) | 58% ( 7) | | | 77% (24) |
| Missing | 0 | 0 | | | 0 |
| **Serious AE** | | | | | |
| Yes | 5% ( 1) | 0% ( 0) | 1.000 | Inf (0.02, Inf) | 3% ( 1) |
| No | 95% (18) | 100% (12) | | | 97% (30) |
| Missing | 0 | 0 | | | 0 |
| **System Organ Class** | | | | | |
| Cardiac disorders | 5% ( 1) | 0% ( 0) | 0.217 | 0.62 (0.35, 0.80) | 3% ( 1) |
| Gastrointestinal disorders | 11% ( 2) | 8% ( 1) | | | 10% ( 3) |
| General disorders and administration site conditions | 5% ( 1) | 0% ( 0) | | | 3% ( 1) |
| Infections and infestations | 47% ( 9) | 17% ( 2) | | | 35% (11) |
| Injury, poisoning and procedural complications | 5% ( 1) | 0% ( 0) | | | 3% ( 1) |
| Investigations | 21% ( 4) | 42% ( 5) | | | 29% ( 9) |
| Nervous system disorders | 0% ( 0) | 8% ( 1) | | | 3% ( 1) |
| Psychiatric disorders | 5% ( 1) | 0% ( 0) | | | 3% ( 1) |
| Respiratory, thoracic and mediastinal disorders | 0% ( 0) | 17% ( 2) | | | 6% ( 2) |
| Vascular disorders | 0% ( 0) | 8% ( 1) | | | 3% ( 1) |
| Missing | 0 | 0 | | | 0 |
| **PreferredTerm** | | | | | |
| Abdominal pain upper | 5% ( 1) | 0% ( 0) | 0.224 | 0.97 (0.93, 0.98) | 3% ( 1) |
| Activated partial thromboplastin time prolonged | 5% ( 1) | 0% ( 0) | | | 3% ( 1) |
| Alanine aminotransferase increased | 5% ( 1) | 0% ( 0) | | | 3% ( 1) |
| Angina pectoris | 5% ( 1) | 0% ( 0) | | | 3% ( 1) |
| Arthropod sting | 5% ( 1) | 0% ( 0) | | | 3% ( 1) |
| Aspartate aminotransferase increased | 5% ( 1) | 0% ( 0) | | | 3% ( 1) |
| Coagulation test abnormal | 0% ( 0) | 8% ( 1) | | | 3% ( 1) |
| Colitis ulcerative | 5% ( 1) | 0% ( 0) | | | 3% ( 1) |
| COVID-19 | 5% ( 1) | 0% ( 0) | | | 3% ( 1) |
| Depressed mood | 5% ( 1) | 0% ( 0) | | | 3% ( 1) |
| Dyspnoea | 0% ( 0) | 8% ( 1) | | | 3% ( 1) |
| Gastrointestinal infection | 5% ( 1) | 0% ( 0) | | | 3% ( 1) |
| Haemorrhoids thrombosed | 0% ( 0) | 8% ( 1) | | | 3% ( 1) |
| Hypoaesthesia | 0% ( 0) | 8% ( 1) | | | 3% ( 1) |
| Influenza | 0% ( 0) | 8% ( 1) | | | 3% ( 1) |
| Influenza like illness | 11% ( 2) | 0% ( 0) | | | 6% ( 2) |
| International normalised ratio abnormal | 0% ( 0) | 17% ( 2) | | | 6% ( 2) |
| International normalised ratio increased | 5% ( 1) | 0% ( 0) | | | 3% ( 1) |
| Lipase increased | 0% ( 0) | 8% ( 1) | | | 3% ( 1) |
| Lymph node palpable | 0% ( 0) | 8% ( 1) | | | 3% ( 1) |
| Nasopharyngitis | 26% ( 5) | 0% ( 0) | | | 16% ( 5) |
| Orchitis | 0% ( 0) | 8% ( 1) | | | 3% ( 1) |
| Periodontitis | 5% ( 1) | 0% ( 0) | | | 3% ( 1) |
| Throat tightness | 0% ( 0) | 8% ( 1) | | | 3% ( 1) |
| Thrombophlebitis | 0% ( 0) | 8% ( 1) | | | 3% ( 1) |
| Missing | 0 | 0 | | | 0 |

NaN: Not a number.

**Supplementary Table 10:** FACIT scores of study participants.

| | | Sequence A | | Sequence B | |
|---|---|---|---|---|---|
| | n | Mean | SD | Mean | SD |
| **V2** | 14 | 18.07 | 8.38 | 20.31 | 7.26 |
| **V4** | 14 | 22.07 | 12.12 | 22.75 | 8.32 |
| **V5** | 14 | 23.79 | 11.18 | 22.19 | 10.26 |
| **V6** | 14 | 24.36 | 11.80 | 23.25 | 11.17 |
| **V7** | 13 | 25.54 | 12.80 | 25.00 | 12.23 |
| **V8** | 13 | 26.23 | 14.30 | 24.50 | 14.08 |
| **V10** | 13 | 25.62 | 12.79 | 25.56 | 13.36 |
| **V11** | 12 | 24.75 | 13.34 | 28.80 | 12.03 |
| **V12** | 12 | 26.50 | 10.72 | 29.13 | 11.61 |
| **V13** | 13 | 28.69 | 12.65 | 28.12 | 13.84 |
| **V14** | 13 | 28.62 | 13.31 | 28.94 | 13.22 |

SD: Standard deviation.

**Supplementary Table 11:** Bell scores of study participants.

| | | Sequence A | | Sequence B | |
|---|---|---|---|---|---|
| | n | Mean | SD | Mean | SD |
| **V2** | 14 | 31.43 | 9.49 | 38.12 | 16.01 |
| **V4** | 14 | 38.57 | 16.10 | 41.25 | 17.84 |
| **V5** | 14 | 38.57 | 17.48 | 37.50 | 17.70 |
| **V6** | 14 | 43.57 | 19.46 | 40.62 | 18.43 |
| **V7** | 13 | 40.00 | 19.15 | 41.25 | 20.29 |
| **V8** | 13 | 42.31 | 20.06 | 42.50 | 20.82 |
| **V10** | 13 | 43.08 | 19.74 | 43.75 | 19.96 |
| **V11** | 12 | 45.00 | 20.23 | 47.33 | 21.20 |
| **V12** | 12 | 45.83 | 19.75 | 46.67 | 23.20 |
| **V13** | 13 | 49.23 | 19.77 | 45.62 | 24.49 |
| **V14** | 13 | 50.77 | 21.39 | 48.12 | 23.16 |

SD: Standard deviation.

**Supplementary Table 12:** FSS scores of study participants.

| | | Sequence A | | Sequence B | |
|---|---|---|---|---|---|
| | n | Mean | SD | Mean | SD |
| **V2** | 14 | 6.24 | 1.46 | 20.31 | 7.26 |
| **V4** | 14 | 5.98 | 1.52 | 22.75 | 8.32 |
| **V5** | 14 | 5.79 | 1.62 | 22.19 | 10.26 |
| **V6** | 14 | 5.72 | 1.85 | 23.25 | 11.17 |
| **V7** | 13 | 5.48 | 1.69 | 25.00 | 12.23 |
| **V8** | 13 | 5.54 | 1.73 | 24.50 | 14.08 |
| **V10** | 13 | 5.47 | 1.74 | 25.56 | 13.36 |
| **V11** | 12 | 5.42 | 1.77 | 28.80 | 12.03 |
| **V12** | 12 | 5.26 | 1.77 | 29.13 | 11.61 |
| **V13** | 13 | 5.10 | 1.85 | 28.12 | 13.84 |
| **V14** | 13 | 5.33 | 1.75 | 28.94 | 13.22 |

SD: Standard deviation.

**Supplementary Table 13:** Canadian consensus criteria (CCC) scores of study participants.

| | | Sequence A | | Sequence B | |
|---|---|---|---|---|---|
| | n | Yes | No | Yes | No |
| **V2** | 14 | 86% (12) | 14% ( 2) | 56% ( 9) | 44% ( 7) |
| **V4** | 14 | 64% ( 9) | 36% ( 5) | 38% ( 6) | 62% (10) |
| **V5** | 14 | 71% (10) | 29% ( 4) | 44% ( 7) | 56% ( 9) |
| **V6** | 14 | 64% ( 9) | 36% ( 5) | 38% ( 6) | 62% (10) |
| **V7** | 13 | 46% ( 6) | 54% ( 7) | 50% ( 8) | 50% ( 8) |
| **V8** | 13 | 46% ( 6) | 54% ( 7) | 38% ( 6) | 62% (10) |
| **V10** | 13 | 46% ( 6) | 54% ( 7) | 38% ( 6) | 62% (10) |
| **V11** | 12 | 50% ( 6) | 50% ( 6) | 20% ( 3) | 80% (12) |
| **V12** | 12 | 50% ( 6) | 50% ( 6) | 20% ( 3) | 80% (12) |
| **V13** | 13 | 38% ( 5) | 62% ( 8) | 25% ( 4) | 75% (12) |
| **V14** | 13 | 46% ( 6) | 54% ( 7) | 38% ( 6) | 62% (10) |

**Supplementary Table 14:** Vitality scores of study participants assessed by SF36 questionnaire.

| | | Sequence A | | Sequence B | |
|---|---|---|---|---|---|
| | **n** | **Mean** | **SD** | **Mean** | **SD** |
| **V2** | 14 | 17.50 | 11.39 | 24.06 | 17.05 |
| **V4** | 14 | 19.29 | 12.84 | 26.88 | 17.97 |
| **V5** | 14 | 23.57 | 16.46 | 24.12 | 18.14 |
| **V6** | 14 | 28.21 | 17.39 | 27.19 | 21.29 |
| **V7** | 13 | 30.77 | 21.59 | 25.00 | 20.17 |
| **V8** | 13 | 29.62 | 18.08 | 27.19 | 22.73 |
| **V10** | 13 | 31.54 | 19.19 | 26.25 | 23.06 |
| **V11** | 12 | 29.58 | 21.37 | 32.00 | 21.78 |
| **V12** | 12 | 33.75 | 20.13 | 31.00 | 25.23 |
| **V13** | 13 | 36.54 | 21.74 | 33.44 | 23.29 |
| **V14** | 13 | 36.15 | 21.62 | 37.19 | 24.01 |

SD: Standard deviation.

**Supplementary Table 15:** General Health scores of study participants assessed by SF36 questionnaire.

| | | Sequence A | | Sequence B | |
|---|---|---|---|---|---|
| | **n** | **Mean** | **SD** | **Mean** | **SD** |
| **V2** | 14 | 25.00 | 15.44 | 34.31 | 15.07 |
| **V4** | 14 | 27.29 | 15.49 | 30.00 | 11.83 |
| **V5** | 14 | 28.36 | 14.84 | 27.81 | 10.16 |
| **V6** | 14 | 30.14 | 14.69 | 31.06 | 14.65 |
| **V7** | 13 | 31.54 | 12.48 | 30.75 | 15.39 |
| **V8** | 13 | 35.46 | 22.30 | 30.56 | 17.10 |
| **V10** | 13 | 32.62 | 20.41 | 32.56 | 16.92 |
| **V11** | 12 | 33.50 | 22.11 | 35.40 | 17.52 |
| **V12** | 12 | 37.42 | 19.71 | 34.27 | 18.12 |
| **V13** | 13 | 40.23 | 20.13 | 31.81 | 15.43 |
| **V14** | 13 | 34.31 | 20.76 | 34.44 | 18.44 |

SD: Standard deviation.

**Supplementary Table 16:** Social role functioning scores of study participants assessed by SF36 questionnaire.

| | | Sequence A | | Sequence B | |
|---|---|---|---|---|---|
| | **n** | **Mean** | **SD** | **Mean** | **SD** |
| **V2** | 14 | 33.93 | 22.70 | 28.12 | 21.65 |
| **V4** | 14 | 31.25 | 23.39 | 27.34 | 22.46 |
| **V5** | 14 | 40.18 | 26.03 | 24.22 | 22.11 |
| **V6** | 14 | 39.29 | 28.10 | 28.12 | 19.63 |
| **V7** | 13 | 43.27 | 29.59 | 28.91 | 24.88 |
| **V8** | 13 | 41.35 | 30.36 | 30.47 | 24.99 |
| **V10** | 13 | 40.38 | 29.82 | 31.25 | 29.58 |
| **V11** | 12 | 35.42 | 31.46 | 38.33 | 23.84 |
| **V12** | 12 | 45.83 | 29.84 | 37.50 | 22.66 |
| **V13** | 13 | 44.23 | 36.31 | 36.72 | 27.94 |
| **V14** | 13 | 50.00 | 33.07 | 39.84 | 30.35 |

SD: Standard deviation.

**Supplementary Table 17:** Mental component summary (MCS) scores of study participants assessed by SF36 questionnaire.

| | | Sequence A | | Sequence B | |
|---|---|---|---|---|---|
| | **n** | **Mean** | **SD** | **Mean** | **SD** |
| **V2** | 14 | 32.54 | 9.96 | 37.17 | 9.90 |
| **V4** | 14 | 34.05 | 8.84 | 36.76 | 8.88 |
| **V5** | 14 | 34.58 | 11.16 | 35.64 | 9.92 |
| **V6** | 14 | 36.90 | 11.21 | 36.56 | 8.37 |
| **V7** | 13 | 37.45 | 12.67 | 35.64 | 8.47 |
| **V8** | 13 | 36.17 | 11.71 | 38.13 | 9.11 |
| **V10** | 13 | 38.44 | 12.50 | 37.72 | 10.93 |
| **V11** | 12 | 35.80 | 12.13 | 37.13 | 10.19 |
| **V12** | 12 | 40.39 | 10.17 | 38.86 | 9.40 |
| **V13** | 13 | 39.78 | 13.54 | 38.32 | 10.35 |
| **V14** | 13 | 39.15 | 12.47 | 41.63 | 9.12 |

SD: Standard deviation.

**Supplementary Table 18:** Physical function scores of study participants assessed by SF36 questionnaire.

| | | Sequence A | | Sequence B | |
|---|---|---|---|---|---|
| | **n** | **Mean** | **SD** | **Mean** | **SD** |
| **V2** | 14 | 46.79 | 21.09 | 48.75 | 22.40 |
| **V4** | 14 | 49.64 | 22.40 | 55.31 | 28.31 |
| **V5** | 14 | 52.50 | 23.43 | 54.38 | 27.44 |
| **V6** | 14 | 52.86 | 24.86 | 57.19 | 28.40 |
| **V7** | 13 | 52.31 | 24.72 | 58.75 | 29.24 |
| **V8** | 13 | 53.85 | 25.75 | 55.31 | 28.08 |
| **V10** | 13 | 54.62 | 24.70 | 58.12 | 27.92 |
| **V11** | 12 | 53.75 | 26.55 | 63.00 | 26.17 |
| **V12** | 12 | 54.58 | 26.06 | 66.67 | 26.97 |
| **V13** | 13 | 53.08 | 25.21 | 62.50 | 26.83 |
| **V14** | 13 | 57.69 | 27.74 | 63.44 | 25.54 |

SD: Standard deviation.

**Supplementary Table 19:** Physical role function scores of study participants assessed by SF36 questionnaire.

| | | Sequence A | | Sequence B | |
|---|---|---|---|---|---|
| | **n** | **Mean** | **SD** | **Mean** | **SD** |
| **V2** | 14 | 7.14 | 26.73 | 4.69 | 13.60 |
| **V4** | 14 | 8.93 | 27.05 | 3.12 | 12.50 |
| **V5** | 14 | 12.50 | 27.30 | 0.00 | 0.00 |
| **V6** | 14 | 12.50 | 27.30 | 6.25 | 14.43 |
| **V7** | 13 | 9.62 | 28.02 | 9.38 | 25.62 |
| **V8** | 13 | 11.54 | 28.17 | 12.50 | 30.28 |
| **V10** | 13 | 15.38 | 33.13 | 9.38 | 27.20 |
| **V11** | 12 | 10.42 | 29.11 | 16.67 | 34.93 |
| **V12** | 12 | 10.42 | 29.11 | 21.67 | 32.55 |
| **V13** | 13 | 19.23 | 38.40 | 26.56 | 37.05 |
| **V14** | 13 | 25.00 | 43.30 | 23.44 | 30.91 |

SD: Standard deviation.

**Supplementary Table 20:** Emotional role functioning scores of study participants assessed by SF36 questionnaire.

| | | Sequence A | | Sequence B | |
|---|---|---|---|---|---|
| | n | Mean | SD | Mean | SD |
| **V2** | 14 | 40.48 | 43.71 | 67.29 | 42.93 |
| **V4** | 14 | 57.14 | 42.22 | 70.83 | 41.94 |
| **V5** | 14 | 45.24 | 44.54 | 70.83 | 41.94 |
| **V6** | 14 | 57.14 | 44.20 | 66.66 | 40.37 |
| **V7** | 13 | 58.97 | 43.36 | 70.83 | 41.94 |
| **V8** | 13 | 56.41 | 45.92 | 72.91 | 38.91 |
| **V10** | 13 | 64.10 | 48.04 | 72.92 | 44.25 |
| **V11** | 12 | 52.78 | 50.17 | 64.44 | 40.76 |
| **V12** | 12 | 63.89 | 45.97 | 82.22 | 35.34 |
| **V13** | 13 | 58.97 | 49.35 | 70.83 | 41.94 |
| **V14** | 13 | 58.97 | 49.35 | 77.08 | 35.94 |

SD: Standard deviation.

**Supplementary Table 21:** Emotional well-being scores of study participants assessed by SF36 questionnaire.

| | | Sequence A | | Sequence B | |
|---|---|---|---|---|---|
| | n | Mean | SD | Mean | SD |
| **V2** | 14 | 64.00 | 11.42 | 68.00 | 12.90 |
| **V4** | 14 | 64.29 | 12.00 | 68.75 | 12.15 |
| **V5** | 14 | 64.86 | 11.22 | 67.50 | 12.64 |
| **V6** | 14 | 66.29 | 14.86 | 70.25 | 13.38 |
| **V7** | 13 | 63.69 | 13.51 | 67.00 | 13.27 |
| **V8** | 13 | 63.08 | 14.44 | 71.50 | 13.38 |
| **V10** | 13 | 67.69 | 14.46 | 72.50 | 13.14 |
| **V11** | 12 | 67.33 | 13.63 | 68.80 | 14.12 |
| **V12** | 12 | 69.33 | 11.36 | 69.87 | 11.80 |
| **V13** | 13 | 68.62 | 11.18 | 71.25 | 13.32 |
| **V14** | 13 | 67.38 | 14.41 | 73.75 | 14.01 |

SD: Standard deviation.

**Supplementary Table 22:** Physical pain scores of study participants assessed by SF36 questionnaire.

| | | Sequence A | | Sequence B | |
|---|---|---|---|---|---|
| | n | Mean | SD | Mean | SD |
| **V2** | 14 | 33.79 | 23.05 | 47.12 | 23.81 |
| **V4** | 14 | 40.00 | 24.07 | 49.06 | 27.04 |
| **V5** | 14 | 37.75 | 20.84 | 48.12 | 29.44 |
| **V6** | 14 | 40.79 | 27.98 | 48.75 | 23.56 |
| **V7** | 13 | 43.85 | 22.34 | 49.50 | 24.57 |
| **V8** | 13 | 43.31 | 27.44 | 49.75 | 28.36 |
| **V10** | 13 | 44.92 | 25.11 | 55.44 | 26.35 |
| **V11** | 12 | 44.83 | 29.15 | 55.33 | 27.38 |
| **V12** | 12 | 47.58 | 23.78 | 58.13 | 23.52 |
| **V13** | 13 | 49.23 | 24.66 | 55.44 | 28.87 |
| **V14** | 13 | 54.23 | 30.93 | 58.88 | 23.46 |

SD: Standard deviation.

**Supplementary Table 23:** Physical component summary (PCS) scores of study participants assessed by SF36 questionnaire.

| | | Sequence A | | Sequence B | |
|---|---|---|---|---|---|
| | **n** | **Mean** | **SD** | **Mean** | **SD** |
| **V2** | 14 | 29.78 | 6.97 | 30.96 | 6.44 |
| **V4** | 14 | 30.59 | 7.44 | 31.77 | 7.85 |
| **V5** | 14 | 31.72 | 6.48 | 31.18 | 7.88 |
| **V6** | 14 | 31.69 | 7.72 | 32.49 | 7.76 |
| **V7** | 13 | 31.99 | 6.92 | 33.50 | 8.72 |
| **V8** | 13 | 33.20 | 8.22 | 32.40 | 8.20 |
| **V10** | 13 | 32.74 | 8.48 | 33.40 | 7.43 |
| **V11** | 12 | 33.40 | 7.86 | 35.97 | 8.20 |
| **V12** | 12 | 32.44 | 8.40 | 36.37 | 7.49 |
| **V13** | 13 | 34.86 | 8.74 | 35.89 | 9.94 |
| **V14** | 13 | 34.90 | 11.47 | 36.11 | 8.17 |

SD: Standard deviation.

**Supplementary Table 24:** Chalder Fatigue scale scores of study participants.

| | | Sequence A | | Sequence B | |
|---|---|---|---|---|---|
| | **n** | **Mean** | **SD** | **Mean** | **SD** |
| **V2** | 14 | 9.93 | 2.40 | 9.75 | 1.39 |
| **V4** | 14 | 9.21 | 3.33 | 9.00 | 2.16 |
| **V5** | 14 | 8.50 | 3.80 | 7.94 | 3.30 |
| **V6** | 14 | 8.71 | 3.77 | 7.38 | 3.88 |
| **V7** | 13 | 8.62 | 2.87 | 7.44 | 3.71 |
| **V8** | 13 | 8.15 | 3.67 | 6.75 | 4.37 |
| **V10** | 13 | 8.15 | 3.74 | 6.44 | 4.00 |
| **V11** | 12 | 8.00 | 3.91 | 5.80 | 3.84 |
| **V12** | 12 | 7.83 | 3.93 | 5.60 | 4.22 |
| **V13** | 13 | 7.15 | 3.93 | 5.44 | 4.52 |
| **V14** | 13 | 7.54 | 3.64 | 5.31 | 4.27 |

SD: Standard deviation.

**Supplementary Table 25:** Walking distance of study participants in meters assessed by 6MWT.

| | | Sequence A | | Sequence B | |
|---|---|---|---|---|---|
| | **n** | **Mean** | **SD** | **Mean** | **SD** |
| **V2** | 14 | 307.64 | 114.56 | 335.38 | 150.92 |
| **V5** | 14 | 325.93 | 139.48 | 351.38 | 182.81 |
| **V7** | 14 | 312.21 | 167.72 | 326.56 | 185.71 |
| **V8** | 13 | 338.77 | 160.82 | 356.38 | 174.77 |
| **V11** | 13 | 289.38 | 167.76 | 375.81 | 182.90 |
| **V13** | 13 | 359.08 | 131.61 | 358.06 | 198.88 |
| **V14** | 13 | 339.85 | 168.03 | 393.06 | 173.33 |

SD: Standard deviation.

**Supplementary Table 26:** Exertion of study participants assessed by 6MWT.

| | | Sequence A | | Sequence B | |
|---|---|---|---|---|---|
| | **n** | **Mean** | **SD** | **Mean** | **SD** |
| **V2** | 13 | 13.85 | 3.13 | 13.25 | 3.70 |
| **V5** | 14 | 12.57 | 2.90 | 12.93 | 3.52 |
| **V7** | 12 | 13.08 | 1.31 | 12.71 | 4.38 |
| **V8** | 13 | 13.15 | 2.15 | 12.64 | 3.00 |
| **V11** | 12 | 12.58 | 3.34 | 12.21 | 3.49 |
| **V13** | 13 | 12.46 | 3.28 | 11.85 | 3.08 |
| **V14** | 12 | 12.50 | 3.63 | 11.21 | 3.31 |

SD: Standard deviation.

**Supplementary Table 27:** Dyspnoea of study participants assessed by 6MWT.

| | | Sequence A | | Sequence B | |
|---|---|---|---|---|---|
| | **n** | **Mean** | **SD** | **Mean** | **SD** |
| **V2** | 13 | 1.15 | 1.86 | 1.44 | 1.90 |
| **V5** | 14 | 1.21 | 2.04 | 1.43 | 2.47 |
| **V7** | 12 | 0.67 | 1.07 | 1.21 | 2.78 |
| **V8** | 13 | 1.15 | 2.15 | 0.96 | 1.97 |
| **V11** | 12 | 0.92 | 1.38 | 1.21 | 2.15 |
| **V13** | 13 | 0.81 | 1.35 | 1.23 | 1.59 |
| **V14** | 12 | 1.00 | 1.71 | 0.32 | 0.42 |

SD: Standard deviation.

**Supplementary Table 28:** DSQ-PEM scores of study participants.

| | | Sequence A | | Sequence B | |
|---|---|---|---|---|---|
| | **n** | **Mean** | **SD** | **Mean** | **SD** |
| **V2** | 14 | 29.64 | 8.54 | 29.38 | 11.23 |
| **V4** | 14 | 31.57 | 9.44 | 29.56 | 11.24 |
| **V5** | 14 | 31.50 | 8.61 | 30.19 | 11.08 |
| **V6** | 14 | 30.50 | 9.94 | 29.75 | 11.58 |
| **V7** | 13 | 29.23 | 10.67 | 28.88 | 12.15 |
| **V8** | 13 | 29.23 | 12.45 | 29.81 | 12.17 |
| **V10** | 13 | 29.46 | 11.20 | 27.94 | 12.55 |
| **V11** | 12 | 28.33 | 11.73 | 26.27 | 11.30 |
| **V12** | 12 | 27.42 | 12.04 | 24.47 | 12.59 |
| **V13** | 13 | 25.23 | 11.57 | 26.38 | 12.25 |
| **V14** | 13 | 26.15 | 12.36 | 23.94 | 12.36 |

SD: Standard deviation.

**Supplementary Table 29:** Positive and negative chronotropic fAAbs of study participants.

| | | Positive chronotropic | | Negative chronotropic | |
|---|---|---|---|---|---|
| Participant | Visit | Positive/Negative | Value | Positive/Negative | Value |
| ER01 | V1 | Positive | 3·83 | Positive | -4·17 |
| | V5 | Negative | 0·00 | Negative | 0·00 |
| | V7 | Negative | -0·33 | Negative | -0·33 |
| | V11 | Negative | 0·00 | Negative | 0·17 |
| | V13 | Positive | 5·17 | Positive | -4·83 |
| ER02 | V1 | Positive | 4·84 | Positive | -4·33 |
| | V5 | Negative | 0·17 | Negative | -0·17 |
| | V7 | Negative | 0·00 | Negative | 0·33 |
| | V11 | Negative | 0·00 | Negative | -0·17 |
| | V13 | Negative | 0·00 | Negative | -0·17 |
| ER03 | V1 | Positive | 4·17 | Positive | -3·00 |
| | V5 | Positive | 6·00 | Positive | -7·67 |
| | V7 | Positive | 3·00 | Positive | -2·50 |
| | V11 | Negative | 0·50 | Negative | -0·83 |
| | V13 | Negative | 0·17 | Negative | 0·67 |
| ER04 | V1 | Positive | 2·17 | Positive | -2·00 |
| | V5 | Positive | 5·00 | Positive | -4·33 |
| | V7 | Positive | 4·67 | Positive | -3·67 |
| | V11 | Negative | 0·17 | Negative | -0·17 |
| | V13 | Negative | 0·00 | Negative | -0·33 |
| ER05 | V1 | Positive | 4·83 | Positive | -5·17 |
| | V5 | Negative | 0·00 | Negative | 0·00 |
| | V7 | Negative | 0·00 | Negative | 0·00 |
| | V11 | Negative | 0·50 | Negative | -0·17 |
| | V13 | Negative | 0·00 | Negative | -0·68 |
| ER06 | V1 | Positive | 4·50 | Positive | -5·00 |
| | V5 | Negative | 0·17 | Negative | -0·33 |
| | V7 | Negative | 0·17 | Negative | 0·17 |
| | V11 | Negative | 0·17 | Negative | -0·33 |
| | V13 | Negative | -0·17 | Negative | 0·00 |
| ER07 | V1 | Positive | 3·50 | Positive | -5·00 |
| | V5 | Negative | 0·17 | Negative | 0·00 |
| | V7 | Negative | 0·17 | Negative | -0·17 |
| | V11 | Negative | -0·50 | Negative | 0·33 |
| | V13 | Negative | -0·17 | Negative | 0·17 |
| ER09 | V1 | Positive | 4·67 | Positive | -5·00 |
| | V5 | Positive | 5·18 | Positive | -5·00 |
| | V7 | Positive | 4·17 | Positive | -6·33 |
| | V11 | Negative | 0·00 | Negative | 0·00 |
| | V13 | Negative | 0·33 | Negative | 0·50 |
| ER10 | V1 | Positive | 5·67 | Positive | -5·00 |
| | V5 | Positive | 4·67 | Positive | -4·00 |
| | V7 | Positive | 3·83 | Positive | -3·00 |
| | V11 | Negative | -0·50 | Negative | 0·17 |
| | V13 | Negative | -0·17 | Negative | -0·17 |
| ER11 | V1 | Positive | 4·50 | Positive | -4·83 |
| | V5 | Positive | 4·83 | Positive | -4·00 |
| | V7 | Positive | 5·17 | Positive | -3·67 |
| | V11 | Negative | 0·50 | Negative | 0·50 |
| | V13 | Negative | 0·17 | Negative | 0·17 |
| ER12 | V1 | Positive | 3·83 | Positive | -3·50 |
| | V5 | Negative | 0·33 | Negative | 0·67 |
| | V7 | Negative | 0·17 | Negative | -0·17 |
| | V11 | Negative | -0·17 | Negative | 0·17 |
| | V13 | Negative | 0·33 | Negative | -0·50 |
| ER13 | V1 | Positive | 2·33 | Positive | -3·00 |
| | V5 | Negative | 0·00 | Negative | 0·17 |
| | V7 | Negative | 0·17 | Negative | -0·17 |
| | V11 | Negative | -0·33 | Negative | 0·33 |
| | V13 | Positive | 3·17 | Negative | -0·17 |
| ER15 | V1 | Positive | 4·00 | Positive | -3·00 |
| | V5 | Positive | 3·50 | Positive | -3·00 |
| | V7 | Positive | 3·33 | Positive | -2·33 |
| | V11 | Negative | 0·00 | Negative | 0·67 |
| | V13 | Negative | 0·17 | Negative | 0·17 |
| ER16 | V1 | Positive | 5·17 | Positive | -3·50 |
| | V5 | Positive | 4·50 | Positive | -4·50 |
| | V7 | Positive | 4·83 | Positive | -3·50 |

| | | | | | |
|---|---|---|---|---|---|
| | V11 | Negative | 0·00 | Negative | -0·17 |
| | V13 | Negative | 0·17 | Negative | 0·17 |
| ER17 | V1 | Positive | 5·17 | Positive | -3·50 |
| | V5 | Negative | 0·17 | Negative | -0·17 |
| | V7 | Negative | -0·17 | Negative | 0·17 |
| | V11 | Negative | -0·50 | Negative | -0·17 |
| | V13 | Negative | 0·17 | Negative | -0·17 |
| ER18 | V1 | Positive | 6·17 | Positive | -4·83 |
| | V5 | Negative | 0·17 | Negative | -0·33 |
| | V7 | Negative | 0·17 | Negative | 0·50 |
| | V11 | Negative | 0·33 | Negative | -0·17 |
| | V13 | Negative | -0·33 | Negative | 0·67 |
| ER20 | V1 | Positive | 5·64 | Positive | -2·17 |
| | V5 | Positive | 3·67 | Positive | -2·00 |
| | V7 | Positive | 4·17 | Positive | -3·50 |
| | V11 | Negative | 0·00 | Negative | -0·33 |
| | V13 | Negative | -0·33 | Negative | -0·17 |
| ER21 | V1 | Positive | 6·00 | Positive | -6·17 |
| | V5 | Negative | 0·17 | Negative | 0·33 |
| | V7 | Negative | 0·83 | Negative | 0·33 |
| | V11 | Negative | 0·33 | Negative | -0·33 |
| | V13 | Negative | 0·00 | Negative | 0·00 |
| ER23 | V1 | Positive | 4·33 | Positive | -4·83 |
| | V5 | Negative | -0·17 | Negative | 0·17 |
| | V7 | Negative | 0·00 | Negative | 0·17 |
| | V11 | Negative | 0·17 | Negative | 0·00 |
| | V13 | Negative | 0·33 | Negative | 0·00 |
| ER24 | V1 | Positive | 6·17 | Positive | -5·00 |
| | V5 | Negative | -0·17 | Negative | 0·33 |
| | V7 | Negative | 0·00 | Negative | 0·17 |
| | V11 | Negative | 0·17 | Negative | 0·33 |
| | V13 | Negative | 0·00 | Negative | 0·17 |
| ER27 | V1 | Positive | 4·00 | Positive | -3·00 |
| | V5 | Negative | -0·33 | Negative | 0·33 |
| | V7 | Negative | -0·33 | Negative | 0·33 |
| | V11 | Negative | -0·33 | Negative | -0·67 |
| | V13 | Negative | -0·17 | Negative | 0·00 |
| ER28 | V1 | Positive | 3·50 | Positive | -4·33 |
| | V5 | Negative | 0·17 | Negative | 0·00 |
| | V7 | Negative | 0·67 | Negative | 0·00 |
| ER29 | V1 | Positive | 6·33 | Positive | -3·00 |
| | V5 | Positive | 5·83 | Positive | -5·00 |
| | V7 | Positive | 5·33 | Positive | -3·50 |
| | V11 | Negative | 0·17 | Negative | 0·33 |
| | V13 | Negative | -0·33 | Negative | -0·33 |
| ER31 | V1 | Positive | 4·67 | Positive | -4·33 |
| | V5 | Positive | 3·17 | Positive | -1·83 |
| | V7 | Positive | 5·17 | Positive | -5·33 |
| | V11 | Negative | -0·50 | Negative | 0·17 |
| | V13 | Negative | -0·17 | Negative | 0·00 |
| ER32 | V1 | Positive | 6·83 | Positive | -6·00 |
| | V5 | Positive | 6·00 | Positive | -5·17 |
| | V7 | Positive | 5·50 | Positive | -4·00 |
| | V11 | Negative | 0·17 | Negative | -0·17 |
| | V13 | Negative | -0·33 | Negative | -0·17 |
| ER33 | V1 | Positive | 5·33 | Positive | -4·67 |
| | V5 | Negative | -0·33 | Negative | 0·00 |
| | V7 | Negative | -0·67 | Negative | -0·17 |
| | V13 | Negative | 0·00 | Positive | -1·83 |
| ER34 | V1 | Positive | 3·00 | Positive | -4·00 |
| | V5 | Positive | 2·83 | Positive | -3·00 |
| | V11 | Negative | 0·17 | Negative | -0·17 |
| | V13 | Negative | 0·33 | Negative | -0·17 |
| ER35 | V1 | Positive | 5·83 | Positive | -5·67 |
| | V5 | Negative | -0·17 | Negative | -0·17 |
| | V7 | Negative | -0·17 | Negative | -0·33 |
| | V11 | Negative | -0·50 | Negative | -0·83 |
| | V13 | Negative | 0·17 | Negative | -0·17 |
| ER36 | V1 | Positive | 4·50 | Positive | -4·33 |
| | V5 | Positive | 4·33 | Positive | -4·33 |
| | V7 | Positive | 3·83 | Positive | -5·17 |
| | V13 | Positive | 4·17 | Positive | -4·50 |
| ER37 | V1 | Positive | 5·00 | Positive | -4·67 |

| | | | | | |
|---|---|---|---|---|---|
| | V5 | Positive | 6·50 | Positive | -5·33 |
| | V7 | Positive | 2·83 | Positive | -3·33 |
| | V11 | Negative | -0·33 | Negative | -0·17 |
| | V13 | Negative | 0·33 | Negative | -0·17 |

ER28 only received one infusion (BC007) at V2 (d0). ER36 only received one infusion (placebo) at V2 (d0).

**Supplementary Table 30:** GPCR-fAAbs of study participants.

| | β2 | | AT1 | | Nociceptor | | α1 | | M2 | | Mas | | ETA | |
|---|---|---|---|---|---|---|---|---|---|---|---|---|---|---|
| | Pos/Neg | Value | Pos/Neg | Value | Pos/Neg | Value | Pos/Neg | Value | Pos/Neg | Value | Pos/Neg | Value | Pos/Neg | Value |
| **ER01** | | | | | | | | | | | | | | |
| V1 | Positive | 1·83 | Negative | 0·17 | Negative | 0·00 | Negative | 0·00 | Positive | -1·83 | Negative | -0·51 | Negative | 0·00 |
| V5 | Negative | 0·00 | Negative | 0·00 | Negative | 0·00 | Negative | 0·00 | Negative | 0·00 | Negative | 0·00 | Negative | 0·00 |
| V7 | Negative | 0·00 | Negative | 0·00 | Negative | 0·00 | Negative | 0·00 | Negative | 0·00 | Negative | 0·00 | Negative | 0·00 |
| V11 | Negative | 0·00 | Negative | 0·00 | Negative | 0·00 | Negative | 0·00 | Negative | 0·00 | Negative | 0·00 | Negative | 0·00 |
| V13 | Positive | 3·17 | Negative | 0·00 | Negative | 0·00 | Negative | 0·00 | Positive | -2·00 | Negative | -0·83 | Negative | 0·00 |
| **ER02** | | | | | | | | | | | | | | |
| V1 | Positive | 2·18 | Negative | 0·49 | Negative | 0·00 | Negative | 0·00 | Positive | -2·67 | Negative | 0·00 | Negative | 0·00 |
| V5 | Negative | 0·00 | Negative | 0·00 | Negative | 0·00 | Negative | 0·00 | Negative | 0·00 | Negative | 0·00 | Negative | 0·00 |
| V7 | Negative | 0·00 | Negative | 0·00 | Negative | 0·00 | Negative | 0·00 | Negative | 0·00 | Negative | 0·00 | Negative | 0·00 |
| V11 | Negative | 0·00 | Negative | 0·00 | Negative | 0·00 | Negative | 0·00 | Negative | 0·00 | Negative | 0·00 | Negative | 0·00 |
| V13 | Negative | 0·00 | Negative | 0·00 | Negative | 0·00 | Negative | 0·00 | Negative | 0·00 | Negative | 0·00 | Negative | 0·00 |
| **ER03** | | | | | | | | | | | | | | |
| V1 | Positive | 2·00 | Negative | 0·17 | Negative | 0·00 | Negative | 0·00 | Positive | -1·17 | Negative | -0·66 | Negative | 0·00 |
| V5 | Positive | 2·50 | Positive | 1·00 | Negative | 0·00 | Negative | 0·00 | Positive | -3·00 | Positive | -1·67 | Negative | 0·00 |
| V7 | Positive | 0·00 | Negative | 0·00 | Negative | 0·00 | Negative | 0·00 | Negative | 0·00 | Negative | 0·00 | Negative | 0·00 |
| V11 | Negative | 0·00 | Negative | 0·00 | Negative | 0·00 | Negative | 0·00 | Negative | 0·00 | Negative | 0·00 | Negative | 0·00 |
| V13 | Negative | 0·00 | Negative | 0·00 | Negative | 0·00 | Negative | 0·00 | Negative | 0·00 | Negative | 0·00 | Negative | 0·00 |
| **ER04** | | | | | | | | | | | | | | |
| V1 | Positive | 2·17 | Negative | 0·00 | Negative | 0·00 | Negative | 0·00 | Positive | -2·00 | Negative | 0·00 | Negative | 0·00 |
| V5 | Positive | 3·00 | Negative | 0·00 | Negative | 0·00 | Negative | 0·00 | Positive | -2·00 | Negative | -0·33 | Negative | 0·00 |
| V7 | Positive | 2·00 | Negative | 0·67 | Negative | 0·00 | Negative | 0·00 | Positive | -2·17 | Negative | 0·00 | Negative | 0·00 |
| V11 | Negative | 0·00 | Negative | 0·00 | Negative | 0·00 | Negative | 0·00 | Negative | 0·00 | Negative | 0·00 | Negative | 0·00 |
| V13 | Negative | 0·00 | Negative | 0·00 | Negative | 0·00 | Negative | 0·00 | Negative | 0·00 | Negative | 0·00 | Negative | 0·00 |
| **ER05** | | | | | | | | | | | | | | |
| V1 | Positive | 2·83 | Negative | 0·00 | Negative | 0·00 | Negative | 0·00 | Positive | -2·50 | Negative | -0·17 | Negative | 0·00 |
| V5 | Negative | 0·00 | Negative | 0·00 | Negative | 0·00 | Negative | 0·00 | Negative | 0·00 | Negative | 0·00 | Negative | 0·00 |
| V7 | Negative | 0·00 | Negative | 0·00 | Negative | 0·00 | Negative | 0·00 | Negative | 0·00 | Negative | 0·00 | Negative | 0·00 |
| V11 | Negative | 0·00 | Negative | 0·00 | Negative | 0·00 | Negative | 0·00 | Negative | 0·00 | Negative | 0·00 | Negative | 0·00 |
| V13 | Negative | 0·00 | Negative | 0·00 | Negative | 0·00 | Negative | 0·00 | Negative | 0·00 | Negative | 0·00 | Negative | 0·00 |
| **ER06** | | | | | | | | | | | | | | |
| V1 | Positive | 2·50 | Negative | 0·00 | Negative | 0·00 | Negative | 0·00 | Positive | -2·67 | Negative | 0·00 | Negative | 0·00 |
| V5 | Negative | 0·00 | Negative | 0·00 | Negative | 0·00 | Negative | 0·00 | Negative | 0·00 | Negative | 0·00 | Negative | 0·00 |
| V7 | Negative | 0·00 | Negative | 0·00 | Negative | 0·00 | Negative | 0·00 | Negative | 0·00 | Negative | 0·00 | Negative | 0·00 |
| V11 | Negative | 0·00 | Negative | 0·00 | Negative | 0·00 | Negative | 0·00 | Negative | 0·00 | Negative | 0·00 | Negative | 0·00 |
| V13 | Negative | 0·00 | Negative | 0·00 | Negative | 0·00 | Negative | 0·00 | Negative | 0·00 | Negative | 0·00 | Negative | 0·00 |
| **ER07** | | | | | | | | | | | | | | |
| V1 | Positive | 1·81 | Negative | 0·00 | Negative | 0·00 | Negative | 0·00 | Positive | -2·67 | Negative | 0·00 | Negative | 0·00 |
| V5 | Negative | 0·00 | Negative | 0·00 | Negative | 0·00 | Negative | 0·00 | Negative | 0·00 | Negative | 0·00 | Negative | 0·00 |
| V7 | Negative | 0·00 | Negative | 0·00 | Negative | 0·00 | Negative | 0·00 | Negative | 0·00 | Negative | 0·00 | Negative | 0·00 |
| V11 | Negative | 0·00 | Negative | 0·00 | Negative | 0·00 | Negative | 0·00 | Negative | 0·00 | Negative | 0·00 | Negative | 0·00 |
| V13 | Negative | 0·00 | Negative | 0·00 | Negative | 0·00 | Negative | 0·00 | Negative | 0·00 | Negative | 0·00 | Negative | 0·00 |
| **ER09** | | | | | | | | | | | | | | |
| V1 | Positive | 2·00 | Negative | 0·67 | Negative | 0·00 | Negative | 0·00 | Positive | -2·33 | Negative | -0·34 | Negative | 0·00 |
| V5 | Positive | 2·00 | Positive | 1·17 | Negative | 0·00 | Negative | 0·00 | Positive | -2·17 | Negative | -0·66 | Negative | 0·00 |
| V7 | Positive | 1·67 | Negative | 0·83 | Negative | 0·00 | Negative | 0·00 | Positive | -3·33 | Negative | 0·00 | Negative | 0·00 |
| V11 | Negative | 0·00 | Negative | 0·00 | Negative | 0·00 | Negative | 0·00 | Negative | 0·00 | Negative | 0·00 | Negative | 0·00 |
| V13 | Negative | 0·00 | Negative | 0·00 | Negative | 0·00 | Negative | 0·00 | Negative | 0·00 | Negative | 0·00 | Negative | 0·00 |
| **ER10** | | | | | | | | | | | | | | |
| V1 | Positive | 3·83 | Negative | 0·18 | Negative | 0·00 | Negative | 0·00 | Positive | -2·33 | Negative | -0·34 | Negative | 0·00 |
| V5 | Positive | 2·67 | Negative | 0·83 | Negative | 0·00 | Negative | 0·00 | Positive | -2·17 | Negative | 0·00 | Negative | 0·00 |
| V7 | Positive | 2·33 | Negative | 0·67 | Negative | 0·00 | Negative | 0·00 | Positive | -1·33 | Negative | -0·34 | Negative | 0·00 |
| V11 | Negative | 0·00 | Negative | 0·00 | Negative | 0·00 | Negative | 0·00 | Negative | 0·00 | Negative | 0·00 | Negative | 0·00 |
| V13 | Negative | 0·00 | Negative | 0·00 | Negative | 0·00 | Negative | 0·00 | Negative | 0·00 | Negative | 0·00 | Negative | 0·00 |
| **ER11** | | | | | | | | | | | | | | |
| V1 | Positive | 2·67 | Negative | 0·00 | Negative | 0·00 | Negative | 0·00 | Positive | -3·50 | Negative | 0·00 | Negative | 0·00 |
| V5 | Positive | 2·83 | Negative | 0·00 | Negative | 0·00 | Negative | 0·00 | Positive | -2·00 | Negative | 0·00 | Negative | 0·00 |
| V7 | Positive | 2·33 | Negative | 0·51 | Negative | 0·00 | Negative | 0·00 | Positive | -1·50 | Negative | 0·00 | Negative | 0·00 |
| V11 | Negative | 0·00 | Negative | 0·00 | Negative | 0·00 | Negative | 0·00 | Negative | 0·00 | Negative | 0·00 | Negative | 0·00 |
| V13 | Negative | 0·00 | Negative | 0·00 | Negative | 0·00 | Negative | 0·00 | Negative | 0·00 | Negative | 0·00 | Negative | 0·00 |
| **ER12** | | | | | | | | | | | | | | |
| V1 | Positive | 3·83 | Negative | 0·00 | Negative | 0·00 | Negative | 0·00 | Positive | -3·50 | Negative | 0·00 | Negative | 0·00 |
| V5 | Negative | 0·00 | Negative | 0·00 | Negative | 0·00 | Negative | 0·00 | Negative | 0·00 | Negative | 0·00 | Negative | 0·00 |
| V7 | Negative | 0·00 | Negative | 0·00 | Negative | 0·00 | Negative | 0·00 | Negative | 0·00 | Negative | 0·00 | Negative | 0·00 |
| V11 | Negative | 0·00 | Negative | 0·00 | Negative | 0·00 | Negative | 0·00 | Negative | 0·00 | Negative | 0·00 | Negative | 0·00 |
| V13 | Negative | 0·00 | Negative | 0·00 | Negative | 0·00 | Negative | 0·00 | Negative | 0·00 | Negative | 0·00 | Negative | 0·00 |
| **ER13** | | | | | | | | | | | | | | |
| V1 | Positive | 2·33 | Negative | 0·00 | Negative | 0·00 | Negative | 0·00 | Positive | -3·00 | Negative | 0·00 | Negative | 0·00 |
| V5 | Negative | 0·00 | Negative | 0·00 | Negative | 0·00 | Negative | 0·00 | Negative | 0·00 | Negative | 0·00 | Negative | 0·00 |
| V7 | Negative | 0·00 | Negative | 0·00 | Negative | 0·00 | Negative | 0·00 | Negative | 0·00 | Negative | 0·00 | Negative | 0·00 |
| V11 | Negative | 0·00 | Negative | 0·00 | Negative | 0·00 | Negative | 0·00 | Negative | 0·00 | Negative | 0·00 | Negative | 0·00 |

| | | | | | | | | | | | | | | |
|---|---|---|---|---|---|---|---|---|---|---|---|---|---|---|
| V13 | Negative | 0·00 | Positive | 3·17 | Negative | 0·00 | Negative | 0·00 | Negative | -0·17 | Negative | 0·00 | Negative | 0·00 |
| **ER15** | | | | | | | | | | | | | | |
| V1 | Positive | 4·00 | Negative | 0·00 | Negative | 0·00 | Negative | 0·00 | Positive | -3·00 | Negative | 0·00 | Negative | 0·00 |
| V5 | Positive | 3·50 | Negative | 0·00 | Negative | 0·00 | Negative | 0·00 | Positive | -3·00 | Negative | 0·00 | Negative | 0·00 |
| V7 | Positive | 3·33 | Negative | 0·00 | Negative | 0·00 | Negative | 0·00 | Positive | -2·33 | Negative | 0·00 | Negative | 0·00 |
| V11 | Negative | 0·00 | Negative | 0·00 | Negative | 0·00 | Negative | 0·00 | Negative | 0·00 | Negative | 0·00 | Negative | 0·00 |
| V13 | Negative | 0·00 | Negative | 0·00 | Negative | 0·00 | Negative | 0·00 | Negative | 0·00 | Negative | 0·00 | Negative | 0·00 |
| **ER16** | | | | | | | | | | | | | | |
| V1 | Positive | 2·17 | Negative | 0·83 | Negative | 0·00 | Negative | 0·00 | Positive | -3·50 | Negative | 0·00 | Negative | 0·00 |
| V5 | Positive | 2·33 | Negative | 0·00 | Negative | 0·00 | Negative | 0·00 | Positive | -2·33 | Negative | 0·00 | Negative | 0·00 |
| V7 | Positive | 2·00 | Negative | 0·83 | Negative | 0·00 | Negative | 0·00 | Positive | -2·00 | Negative | 0·00 | Negative | 0·00 |
| V11 | Negative | 0·00 | Negative | 0·00 | Negative | 0·00 | Negative | 0·00 | Negative | 0·00 | Negative | 0·00 | Negative | 0·00 |
| V13 | Negative | 0·00 | Negative | 0·00 | Negative | 0·00 | Negative | 0·00 | Negative | 0·00 | Negative | 0·00 | Negative | 0·00 |
| **ER17** | | | | | | | | | | | | | | |
| V1 | Positive | 2·33 | Negative | 0·51 | Negative | 0·00 | Negative | 0·00 | Positive | -2·17 | Negative | 0·00 | Negative | 0·00 |
| V5 | Negative | 0·00 | Negative | 0·00 | Negative | 0·00 | Negative | 0·00 | Negative | 0·00 | Negative | 0·00 | Negative | 0·00 |
| V7 | Negative | 0·00 | Negative | 0·00 | Negative | 0·00 | Negative | 0·00 | Negative | 0·00 | Negative | 0·00 | Negative | 0·00 |
| V11 | Negative | 0·00 | Negative | 0·00 | Negative | 0·00 | Negative | 0·00 | Negative | 0·00 | Negative | 0·00 | Negative | 0·00 |
| V13 | Negative | 0·00 | Negative | 0·00 | Negative | 0·00 | Negative | 0·00 | Negative | 0·00 | Negative | 0·00 | Negative | 0·00 |
| **ER18** | | | | | | | | | | | | | | |
| V1 | Positive | 3·00 | Negative | 0·17 | Negative | 0·00 | Negative | 0·00 | Positive | -2·00 | Negative | -0·83 | Negative | 0·00 |
| V5 | Negative | 0·00 | Negative | 0·00 | Negative | 0·00 | Negative | 0·00 | Negative | 0·00 | Negative | 0·00 | Negative | 0·00 |
| V7 | Negative | 0·00 | Negative | 0·00 | Negative | 0·00 | Negative | 0·00 | Negative | 0·00 | Negative | 0·00 | Negative | 0·00 |
| V11 | Negative | 0·00 | Negative | 0·00 | Negative | 0·00 | Negative | 0·00 | Negative | 0·00 | Negative | 0·00 | Negative | 0·00 |
| V13 | Negative | 0·00 | Negative | 0·00 | Negative | 0·00 | Negative | 0·00 | Negative | 0·00 | Negative | 0·00 | Negative | 0·00 |
| **ER20** | | | | | | | | | | | | | | |
| V1 | Positive | 2·33 | Negative | 0·98 | Negative | 0·00 | Negative | 0·00 | Positive | -2·17 | Negative | 0·00 | Negative | 0·00 |
| V5 | Positive | 1·83 | Negative | 0·00 | Negative | 0·00 | Negative | 0·00 | Positive | -2·00 | Negative | 0·00 | Negative | 0·00 |
| V7 | Positive | 2·17 | Negative | 0·00 | Negative | 0·00 | Negative | 0·00 | Positive | -2·00 | Negative | 0·00 | Negative | 0·00 |
| V11 | Negative | 0·00 | Negative | 0·00 | Negative | 0·00 | Negative | 0·00 | Negative | 0·00 | Negative | 0·00 | Negative | 0·00 |
| V13 | Negative | 0·00 | Negative | 0·00 | Negative | 0·00 | Negative | 0·00 | Negative | 0·00 | Negative | 0·00 | Negative | 0·00 |
| **ER21** | | | | | | | | | | | | | | |
| V1 | Positive | 3·50 | Negative | 0·33 | Negative | 0·84 | Negative | 0·00 | Positive | -2·67 | Negative | -0·83 | Negative | 0·00 |
| V5 | Negative | 0·00 | Negative | 0·00 | Negative | 0·00 | Negative | 0·00 | Negative | 0·00 | Negative | 0·00 | Negative | 0·00 |
| V7 | Negative | 0·00 | Negative | 0·00 | Negative | 0·00 | Negative | 0·00 | Negative | 0·00 | Negative | 0·00 | Negative | 0·00 |
| V11 | Negative | 0·00 | Negative | 0·00 | Negative | 0·00 | Negative | 0·00 | Negative | 0·00 | Negative | 0·00 | Negative | 0·00 |
| V13 | Negative | 0·00 | Negative | 0·00 | Negative | 0·00 | Negative | 0·00 | Negative | 0·00 | Negative | 0·00 | Negative | 0·00 |
| **ER23** | | | | | | | | | | | | | | |
| V1 | Positive | 4·33 | Negative | 0·00 | Negative | 0·00 | Negative | 0·00 | Positive | -4·83 | Negative | 0·00 | Negative | 0·00 |
| V5 | Negative | 0·00 | Negative | 0·00 | Negative | 0·00 | Negative | 0·00 | Negative | 0·00 | Negative | 0·00 | Negative | 0·00 |
| V7 | Negative | 0·00 | Negative | 0·00 | Negative | 0·00 | Negative | 0·00 | Negative | 0·00 | Negative | 0·00 | Negative | 0·00 |
| V11 | Negative | 0·00 | Negative | 0·00 | Negative | 0·00 | Negative | 0·00 | Negative | 0·00 | Negative | 0·00 | Negative | 0·00 |
| V13 | Negative | 0·00 | Negative | 0·00 | Negative | 0·00 | Negative | 0·00 | Negative | 0·00 | Negative | 0·00 | Negative | 0·00 |
| **ER24** | | | | | | | | | | | | | | |
| V1 | Positive | 3·33 | Positive | 1·34 | Negative | 0·00 | Negative | 0·00 | Positive | -2·83 | Negative | 0·00 | Negative | 0·00 |
| V5 | Negative | 0·00 | Negative | 0·00 | Negative | 0·00 | Negative | 0·00 | Negative | 0·00 | Negative | 0·00 | Negative | 0·00 |
| V7 | Negative | 0·00 | Negative | 0·00 | Negative | 0·00 | Negative | 0·00 | Negative | 0·00 | Negative | 0·00 | Negative | 0·00 |
| V11 | Negative | 0·00 | Negative | 0·00 | Negative | 0·00 | Negative | 0·00 | Negative | 0·00 | Negative | 0·00 | Negative | 0·00 |
| V13 | Negative | 0·00 | Negative | 0·00 | Negative | 0·00 | Negative | 0·00 | Negative | 0·00 | Negative | 0·00 | Negative | 0·00 |
| **ER27** | | | | | | | | | | | | | | |
| V1 | Positive | 4·00 | Negative | 0·00 | Negative | 0·00 | Negative | 0·00 | Positive | -3·00 | Negative | 0·00 | Negative | 0·00 |
| V5 | Negative | 0·00 | Negative | 0·00 | Negative | 0·00 | Negative | 0·00 | Negative | 0·00 | Negative | 0·00 | Negative | 0·00 |
| V7 | Negative | 0·00 | Negative | 0·00 | Negative | 0·00 | Negative | 0·00 | Negative | 0·00 | Negative | 0·00 | Negative | 0·00 |
| V11 | Negative | 0·00 | Negative | 0·00 | Negative | 0·00 | Negative | 0·00 | Negative | 0·00 | Negative | 0·00 | Negative | 0·00 |
| V13 | Negative | 0·00 | Negative | 0·00 | Negative | 0·00 | Negative | 0·00 | Negative | 0·00 | Negative | 0·00 | Negative | 0·00 |
| **ER28** | | | | | | | | | | | | | | |
| V1 | Positive | 3·50 | Negative | 0·00 | Negative | 0·00 | Negative | 0·00 | Positive | -2·50 | Negative | 0·00 | Negative | 0·00 |
| V5 | Negative | 0·00 | Negative | 0·00 | Negative | 0·00 | Negative | 0·00 | Negative | 0·00 | Negative | 0·00 | Negative | 0·00 |
| V7 | Negative | 0·00 | Negative | 0·00 | Negative | 0·00 | Negative | 0·00 | Negative | 0·00 | Negative | 0·00 | Negative | 0·00 |
| **ER29** | | | | | | | | | | | | | | |
| V1 | Positive | 2·33 | Negative | 0·00 | Negative | 0·00 | Negative | 0·00 | Positive | -3·00 | Negative | 0·00 | Negative | 0·00 |
| V5 | Positive | 2·17 | Positive | 1·49 | Negative | 0·00 | Negative | 0·00 | Positive | -2·67 | Negative | 0·00 | Negative | 0·00 |
| V7 | Positive | 3·00 | Negative | 0·00 | Negative | 0·00 | Negative | 0·00 | Positive | 1·50 | Negative | 0·00 | Negative | 0·00 |
| V11 | Negative | 0·00 | Negative | 0·00 | Negative | 0·00 | Negative | 0·00 | Negative | 0·00 | Negative | 0·00 | Negative | 0·00 |
| V13 | Negative | 0·00 | Negative | 0·00 | Negative | 0·00 | Negative | 0·00 | Negative | 0·00 | Negative | 0·00 | Negative | 0·00 |
| **ER31** | | | | | | | | | | | | | | |
| V1 | Positive | 2·50 | Negative | 0·00 | Negative | 0·00 | Negative | 0·00 | Positive | -2·00 | Negative | 0·00 | Negative | 0·00 |
| V5 | Positive | 3·17 | Negative | 0·00 | Negative | 0·00 | Negative | 0·00 | Positive | -1·83 | Negative | 0·00 | Negative | 0·00 |
| V7 | Positive | 1·67 | Positive | 1·83 | Negative | 0·00 | Negative | 0·00 | Positive | -2·33 | Negative | -0·33 | Negative | 0·00 |
| V11 | Negative | 0·00 | Negative | 0·00 | Negative | 0·00 | Negative | 0·00 | Negative | 0·00 | Negative | 0·00 | Negative | 0·00 |
| V13 | Negative | 0·00 | Negative | 0·00 | Negative | 0·00 | Negative | 0·00 | Negative | 0·00 | Negative | 0·00 | Negative | 0·00 |
| **ER32** | | | | | | | | | | | | | | |
| V1 | Positive | 4·00 | Positive | 1·66 | Negative | 0·00 | Negative | 0·00 | Positive | -3·00 | Negative | 0·00 | Negative | 0·00 |
| V5 | Positive | 1·83 | Positive | 2·34 | Negative | 0·00 | Negative | 0·00 | Positive | -2·67 | Negative | 0·00 | Negative | 0·00 |
| V7 | Positive | 2·17 | Positive | 1·16 | Negative | 0·00 | Negative | 0·00 | Positive | -1·67 | Negative | 0·00 | Negative | 0·00 |
| V11 | Negative | 0·00 | Negative | 0·00 | Negative | 0·00 | Negative | 0·00 | Negative | 0·00 | Negative | 0·00 | Negative | 0·00 |

| | | | | | | | | | | | | | | |
|---|---|---|---|---|---|---|---|---|---|---|---|---|---|---|
| V13 | Negative | 0·00 | Negative | 0·00 | Negative | 0·00 | Negative | 0·00 | Negative | 0·00 | Negative | 0·00 | Negative | 0·00 |
| **ER33** | | | | | | | | | | | | | | |
| V1 | Positive | 2·83 | Negative | 0·00 | Negative | 0·00 | Negative | 0·00 | Positive | -2·33 | Negative | 0·00 | Negative | 0·00 |
| V5 | Negative | 0·00 | Negative | 0·00 | Negative | 0·00 | Negative | 0·00 | Negative | 0·00 | Negative | 0·00 | Negative | 0·00 |
| V7 | Negative | 0·00 | Negative | 0·00 | Negative | 0·00 | Negative | 0·00 | Negative | 0·00 | Negative | 0·00 | Negative | 0·00 |
| V13 | Positive | 0·00 | Negative | 0·00 | Negative | 0·00 | Negative | 0·00 | Positive | -1·83 | Negative | 0·00 | Negative | 0·00 |
| **ER34** | | | | | | | | | | | | | | |
| V1 | Positive | 3·00 | Negative | 0·00 | Negative | 0·00 | Negative | 0·00 | Positive | -4·00 | Negative | 0·00 | Negative | 0·00 |
| V5 | Positive | 2·83 | Negative | 0·00 | Negative | 0·00 | Negative | 0·00 | Positive | -3·00 | Negative | 0·00 | Negative | 0·00 |
| V11 | Negative | 0·00 | Negative | 0·00 | Negative | 0·00 | Negative | 0·00 | Negative | 0·00 | Negative | 0·00 | Negative | 0·00 |
| V13 | Negative | 0·00 | Negative | 0·00 | Negative | 0·00 | Negative | 0·00 | Negative | 0·00 | Negative | 0·00 | Negative | 0·00 |
| **ER35** | | | | | | | | | | | | | | |
| V1 | Positive | 3·67 | Negative | 0·00 | Negative | 0·00 | Negative | 0·00 | Positive | -2·50 | Negative | 0·00 | Negative | 0·00 |
| V5 | Negative | 0·00 | Negative | 0·00 | Negative | 0·00 | Negative | 0·00 | Negative | 0·00 | Negative | 0·00 | Negative | 0·00 |
| V7 | Negative | 0·00 | Negative | 0·00 | Negative | 0·00 | Negative | 0·00 | Negative | 0·00 | Negative | 0·00 | Negative | 0·00 |
| V11 | Negative | 0·00 | Negative | 0·00 | Negative | 0·00 | Negative | 0·00 | Negative | 0·00 | Negative | 0·00 | Negative | 0·00 |
| V13 | Negative | 0·00 | Negative | 0·00 | Negative | 0·00 | Negative | 0·00 | Negative | 0·00 | Negative | 0·00 | Negative | 0·00 |
| **ER36** | | | | | | | | | | | | | | |
| V1 | Positive | 2·00 | Negative | 0·00 | Negative | 0·00 | Negative | 0·00 | Positive | -2·00 | Negative | 0·00 | Negative | 0·00 |
| V5 | Positive | 1·67 | Negative | 0·00 | Negative | 0·00 | Negative | 0·00 | Positive | -2·17 | Negative | 0·00 | Negative | 0·00 |
| V7 | Positive | 1·50 | Negative | 0·00 | Negative | 0·00 | Negative | 0·00 | Positive | -2·33 | Negative | 0·00 | Negative | 0·00 |
| V13 | Positive | 3·00 | Negative | 0·00 | Negative | 0·00 | Negative | 0·00 | Positive | -2·17 | Negative | 0·00 | Negative | 0·00 |
| **ER37** | | | | | | | | | | | | | | |
| V1 | Positive | 2·50 | Negative | 0·00 | Negative | 0·00 | Negative | 0·00 | Positive | -2·33 | Negative | 0·00 | Negative | 0·00 |
| V5 | Positive | 4·17 | Negative | 0·00 | Negative | 0·00 | Negative | 0·00 | Positive | -1·83 | Positive | -1·67 | Negative | 0·00 |
| V7 | Positive | 2·83 | Negative | 0·00 | Negative | 0·00 | Negative | 0·00 | Positive | -3·33 | Negative | 0·00 | Negative | 0·00 |
| V11 | Negative | 0·00 | Negative | 0·00 | Negative | 0·00 | Negative | 0·00 | Negative | 0·00 | Negative | 0·00 | Negative | 0·00 |
| V13 | Negative | 0·00 | Negative | 0·00 | Negative | 0·00 | Negative | 0·00 | Negative | 0·00 | Negative | 0·00 | Negative | 0·00 |

ER28 only received one infusion (BC007) at V2 (d0). ER36 only received one infusion (placebo) at V2 (d0).
